# Tissue-specific and circulatory immune signatures of mucosal inflammation in Crohn’s disease

**DOI:** 10.1101/2025.06.03.25328858

**Authors:** Lucia Ramirez-Navarro, Gareth-Rhys Jones, Tobi Alegbe, Bradley T. Harris, Michelle Strickland, Mennatallah Ghouraba, Jasmin Ostermayer, Noor Wana, May Xueqi Hu, Jason Skelton, Wendy Garri, Biljana Brezina, Charry Queen Caballes, Steven Leonard, Vivek Iyer, Rebecca E. McIntyre, Miles Parkes, Cristina Cotobal Martin, Carl A. Anderson, Tim Raine

**Affiliations:** Wellcome Sanger Institute, Hinxton, CB10 1SA, UK; School of Infection and Immunity, University of Glasgow, Glasgow, G12 8QQ, UK; Addenbrooke’s Hospital, Cambridge CB2 0QQ, UK

**Author notes:** Correspondence C.A.A., T.R. These authors jointly supervised the study (Carl A. Anderson, Tim Raine).

**Keywords:** Inflammatory bowel disease, IBD, single-cell RNA sequencing, Crohn’s disease, tissue-resident memory cells, Trm, ex-Trm, mucosal inflammation, blood, gut

## Abstract

Crohn’s disease (CD) is a heterogeneous disease characterized by chronic inflammation of the gastrointestinal tract driven by an aberrant immune response. To understand the immune mechanisms underlying mucosal inflammation, we performed single-cell RNA sequencing on paired terminal ileum biopsies and blood samples from 125 CD patients. Our findings reveal that the transcriptional profiles of immune cells are primarily shaped by the local tissue microenvironment, with mucosal immune cells undergoing profound inflammation-associated changes, while circulating immune cells largely fail to reflect these signatures. However, a subset of circulating T cells with a tissue-resident memory-like phenotype retained transcriptional hallmarks of mucosal inflammation, mirroring their ileal counterparts with enrichment in cytokine pathways, including interferon gamma and TNF receptor signaling. Altogether, our single-cell atlas provides a high-resolution resource for dissecting the mucosal and circulatory immune landscape of CD and its link to mucosal inflammation.

## Introduction

Crohn’s disease (CD) is a common form of inflammatory bowel disease (IBD) characterised by chronic inflammation of the gastrointestinal tract. While its biological basis is poorly understood, it is thought to be driven by an aberrant immune response against commensal gut bacteria in genetically susceptible individuals^1^. Although treatment options have expanded, substantial heterogeneity in disease location, severity, and progression complicates clinical decision-making. Consequently, there is an urgent need for improved understanding of immunopathogenesis and robust biomarkers to guide therapy and predict mucosal healing.

Routine endoscopic monitoring and mucosal sampling are invasive and impractical for longitudinal assessment of intestinal inflammation, prompting interest in blood-based biomarkers as a minimally invasive and accessible alternative. Given that CD can also present extra-intestinal manifestations, systemic immune signatures may offer relevant insights; however the extent to which they reflect gut-specific immune activity remains unclear.

Previous studies have provided valuable insights into tissue-specific immune variation by comparing the transcriptional profiles of blood and mucosal immune cells in healthy individuals^2–4^. While some studies have analyzed paired blood and tissue samples from IBD patients^5–8^, these have been limited in sample size and have not systematically characterised tissue-specific immune variation across cell types.

To address this gap, we performed single-cell RNA sequencing (scRNA-seq) on paired terminal ileum (TI) biopsies and blood samples from 125 CD patients (Fig. 1a and Supplementary Fig. 1), capturing tissue and circulating immune populations across a spectrum of inflammation (Fig. 1b and Supplementary Table 1). This matched design allowed us to assess how immune cell transcriptomes vary with mucosal inflammation and whether changes in tissue are reflected in circulation.

**Figure 1.**
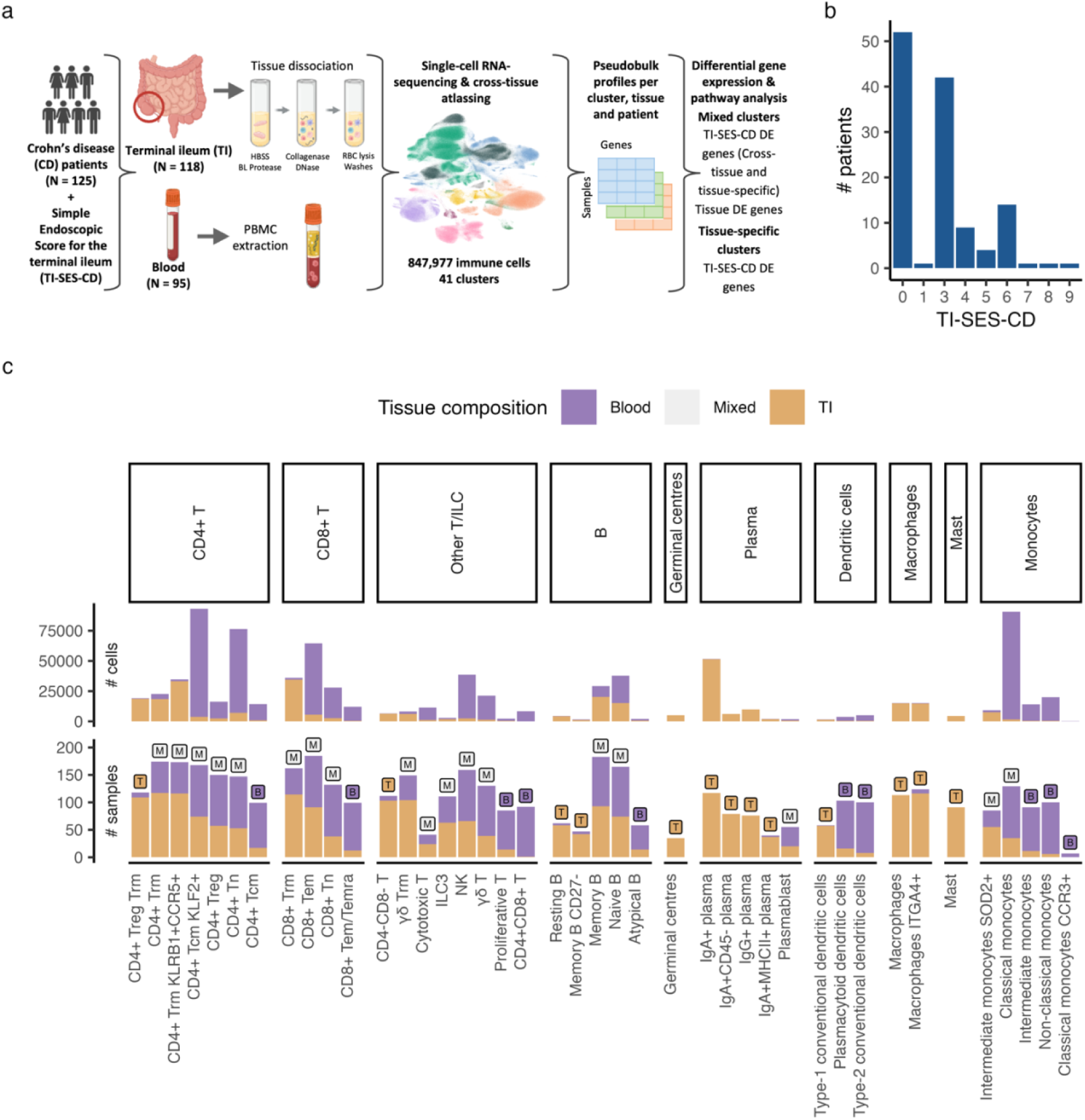
An atlas of mucosal and circulating immune cells in Crohn’s disease patients. a, Study overview. b, Distribution of patients based on the Simple Endoscopic Score for Crohn Disease at the terminal ileum (TI-SES-CD). c, Number of cells (top) and contributing samples (bottom) per immune cell cluster, colored by their tissue of origin (blood or TI). Clusters were classified as tissue specific or mixed based on sample composition: clusters with ≥25% contribution from both tissues were considered mixed. Only samples contributing ≥10 cells to a given cluster were included in the sample count

We found that mucosal immune cells exhibited marked transcriptional changes associated with inflammation severity, involving both shared and cell type-specific responses. In contrast, most circulating immune cells lacked corresponding transcriptional signatures, suggesting that the local tissue environment plays a dominant role in shaping immune states. The key exception was a subset of circulating T cells with a tissue-resident memory (Trm)-like profile, which retained inflammation-associated gene expression resembling their mucosal counterparts. Altogether our single-cell immune atlas provides a valuable resource for dissecting shared and tissue-specific immune transcriptional programs under chronic inflammation and informs future efforts to develop precision diagnostics and therapeutics using blood-based readouts.

## Results

### An atlas of mucosal and circulating immune cells in Crohn’s disease

To characterise the transcriptional profile of mucosal and circulating immune cells in CD, blood and TI biopsies were collected from 125 patients (Fig. 1a). Mucosal inflammation was quantified using the three inflammatory components (ulceration, ulcer size, affected area) of the Simple Endoscopic Score for CD assessed at the TI region (TI-SES-CD) (Fig. 1b and Supplementary Table 1). After quality control, 847,977 cells were analyzed, comprising 537,274 circulating and 310,703 mucosal immune cells. Integration and clustering of cells from both tissues yielded 41 immune subpopulations across three major lineages: B cells, T/ILC cells and myeloid (Supplementary Fig. 2a and Supplementary Table 2). Each cluster was classified based on its tissue distribution at the sample level as either mixed (≥25% contribution from both blood and TI) or tissue-specific (>75% contribution from a single tissue) (Methods), resulting in 18 mixed, 13 TI-specific and 10 blood-specific clusters (Fig. 1c).

Several previously reported immune populations were readily identifiable within the atlas, providing internal validation of our clustering approach. For example, we identified *SOD2*^+^ intermediate monocytes, previously reported in the inflamed gut of CD patients^9^, and now shown to also circulate in peripheral blood (Fig. 1c and Supplementary Fig. 2b). We also resolved a population of *CD27*^−^ memory B cells (Fig. 1c and Supplementary Fig. 2c), originally detected by mass cytometry in IBD patients^10^, but not characterised at the single-cell resolution. These findings highlight the utility of the dataset for both high-resolution mapping and contextualising known immune states.

Within the T cell compartment, we identified multiple Trm subsets, including CD4^+^, CD8^+^, regulatory (Treg) and γδ T cells (Fig. 1c). These populations shared canonical residency signatures, characterized by elevated expression of *ITGAE (CD103), ITGA1 (CD49a), SPRY1, CXCR6, CD101, and CCR9,* and reduced expression of *ITGB1* and *SELL,* relative to other T cells (Supplementary Fig. 2d). γδ Trm cells also showed higher expression of *KLRC2* and *ENTPD1*, markers previously linked with intestinal γδ T cells^11^. Among CD4^+^ Trms, one subset highly expressed *KLRB1 (CD161)* and *CCR5* (Supplementary Fig. 2d), consistent with a pro-inflammatory Trm population described in CD patients^12^. This subset has been reported to produce type 1 inflammatory cytokines independently of TCR ligation and damage adjacent epithelial cells^12^.

Although most Trm cells were derived from the TI, we observed a subset of CD8^+^ and CD4^+^ T cells (including the pro-inflammatory subset) originating from the blood (Fig. 1c and Fig. 2a-b). These circulating cells retained a Trm-like transcriptional signature, marked by elevated expression of residency markers (*ITGAE, ITGA1, CD101, SPRY1, CCR9)* and reduced expression of genes associated with circulating cells (*KLF2*, *S1PR1)* compared to other blood T cells (Fig. 2c). These findings suggest Trm cells can be found in the blood, consistent with Trms that re-entered circulation (ex-Trms).

**Fig. 2.**
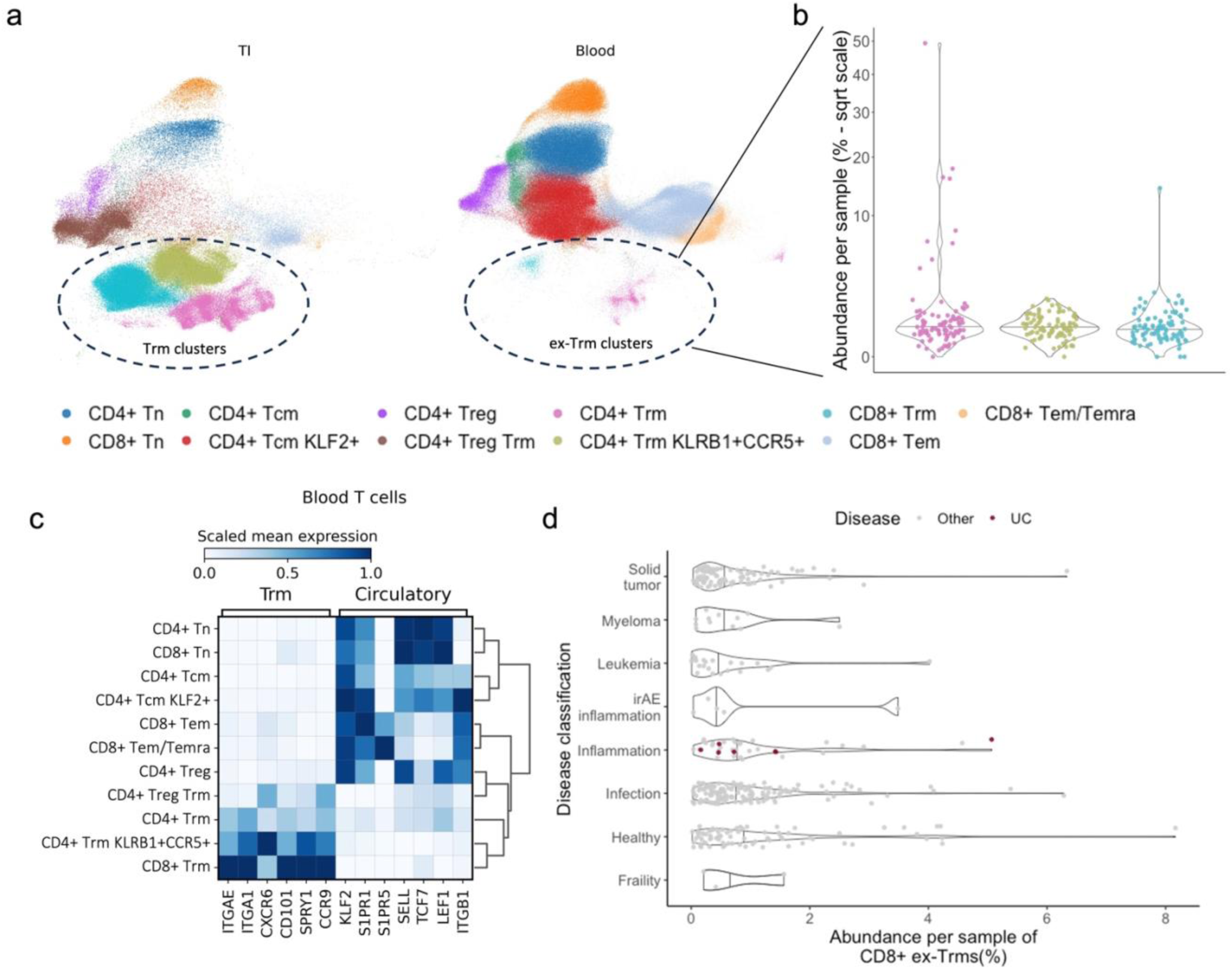
Characterization of circulating ex-Trms in blood. a, Uniform Manifold Approximation and Projection (UMAP) of T cells from the terminal ileum (TI) and blood, colored by cluster identity. Tissue-resident memory (Trm) and circulating ex-Trm clusters are highlighted with dashed circles. b, Per-sample abundance of ex-Trm clusters within circulating T cells. Colors correspond to clusters shown in (a). c, Heatmap showing scaled mean expression levels of genes characteristic of tissue-resident and circulating T cells, comparing ex-Trm clusters to other circulating T-cell populations. d, Abundance of cells predicted as CD8^+^ ex-Trms (confidence > 0.5) across various PBMC datasets and disease conditions in a meta-analysis of CD8^+^ T cells. Disease classifications are derived from the original publication, with ulcerative colitis (UC) samples highlighted in red. For detailed disease-specific abundances, see Supplementary Fig. 4a.

Gut-derived ex-Trms have been described in mouse models^13–15^, where they were shown to exit the intestine and recirculate in response to local activation. In humans, ex-Trms have only been shown to originate from the skin, where Trm-like T cells have been detected in the blood and shown to share transcriptional and clonal features with their skin-resident counterparts^16,17^. While skin-derived ex-Trms typically constitute <1% of CD4⁺ T cells in healthy individuals, we found that the three Trm-like T cell clusters accounted for ∼3% of circulating T cells in our CD cohort (Fig. 2a-b), with a higher proportion of CD4^+^ T cells, consistent with previous observations in skin^17^. All 95 blood samples contained cells from at least one ex-Trm cluster, and 43 samples (45%) had >50 cells across the three clusters, supporting their robust presence in circulation. Ex-Trm abundance was not associated with TI-SES-CD, medication, sex, age or smoking status (Supplementary Fig. 3 and Supplementary Table 3).

To further support the presence of a gut-derived ex-Trm population in blood, we trained a CellTypist model^2,18^ on our dataset and applied it to a meta-analysis of scRNA-seq studies of blood CD8^+^ T cells^19^. This analysis identified a rare CD8^+^ T cell population in both IBD patients and healthy individuals (Fig. 2d and Supplementary Fig. 4a) that displayed a transcriptomic signature similar to the ex-Trms we observed, including expression of the gut homing receptor *CCR9* (Supplementary Fig. 4b). These findings suggest that a blood-based ex-Trm population, although infrequent, is reproducibly identifiable across independent cohorts.

### ex-Trms are the only circulating cells that recapitulate mucosal inflammation

We next asked whether any immune cell populations shared transcriptional signatures of inflammation across both blood and gut. If so, this would suggest that local inflammatory signals may disseminate systemically, indicating that shared effector states can emerge across tissues in response to the degree of local inflammation.

To test this, we performed differential gene expression (DGE) analyses using TI-SES-CD scores as a continuous variable (Methods). For each cluster containing cells from both tissues (mixed clusters), we identified two categories of inflammation-associated genes: (1) genes whose expression correlated with TI-SES-CD in both blood and tissue (cross-tissue), and (2) genes with correlations restricted to a single tissue, detected through a tissue-by-inflammation interaction term (tissue-specific) (Supplementary Table 4). Among the mixed clusters, only CD8^+^ and CD4^+^ Trms, including the pro-inflammatory *KLRB1^+^CCR5^+^* subset, exhibited significant (FDR<0.05) cross-tissue gene expression, with 98 genes in CD4^+^ Trms, 84 in the *KLRB1^+^CCR5^+^* subset, and 3 in CD8^+^ Trms. In contrast, the remaining mixed clusters had no significant cross-tissue genes and contained only tissue-specific associations (Fig. 3a). Within the tissue-specific set, 415 genes were correlated with TI-SES-CD scores exclusively in TI-derived cells, none were specific to circulating cells and 10 genes showed opposing correlations between the two tissues (Supplementary Table 5). The absence of inflammatory signatures in other circulating cells is further supported by the analysis of tissue-specific clusters, where we tested for correlations with TI-SES-CD using only cells from their respective tissue of origin (Methods). Among the 10 blood-specific and 11 ileal-specific clusters tested, only the ileal clusters showed significant associations, with 5,428 distinct genes linked to TI-SES-CD scores (Fig. 3b and Supplementary Table 4). Despite similar sample sizes (Fig. 1c), none of the blood-specific clusters contained genes whose expression was associated with mucosal inflammation.

**Figure 3.**
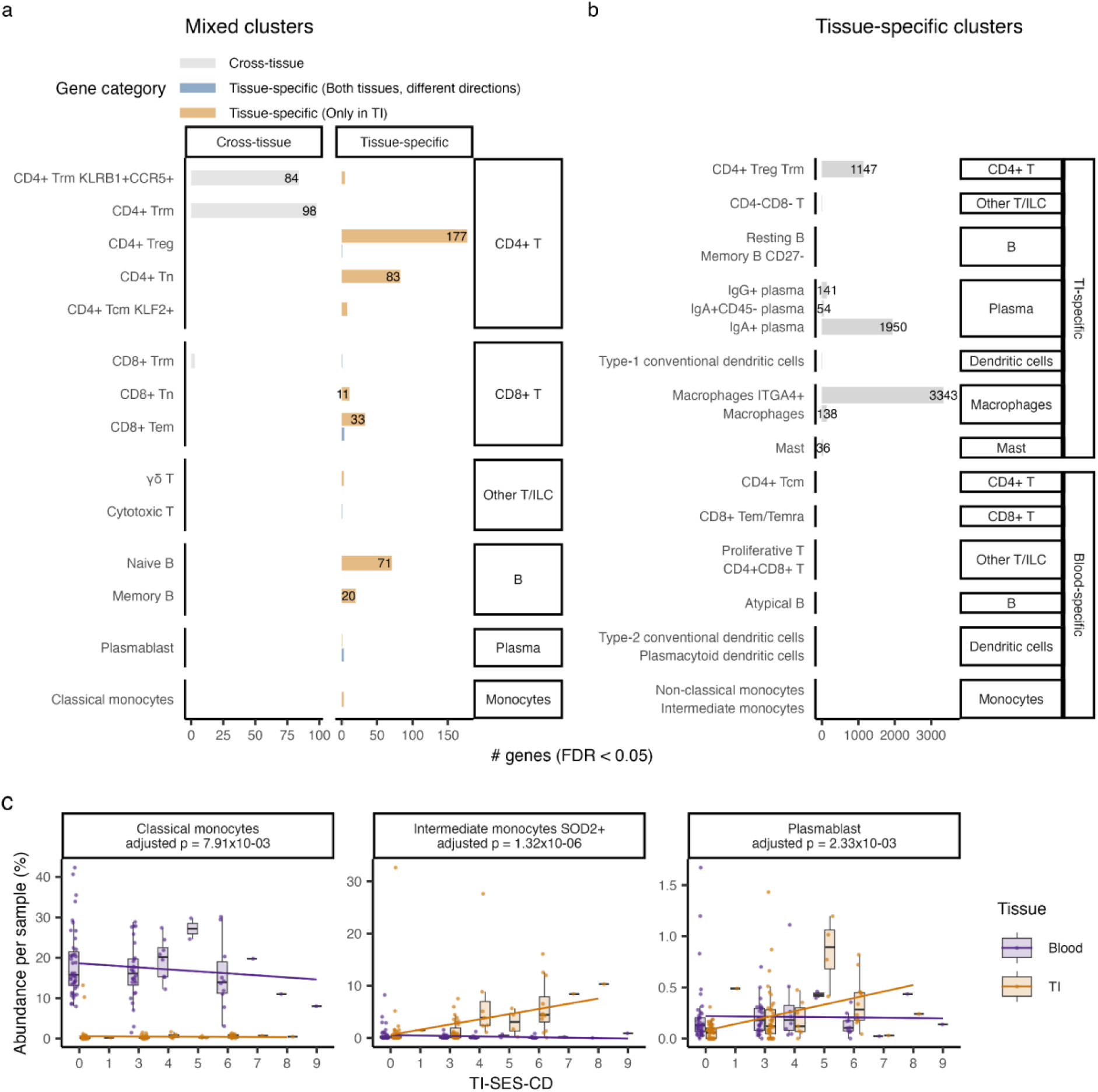
Transcriptional and compositional associations of immune cells with mucosal inflammation. a, Number of differentially expressed (DE) genes (FDR < 0.05) identified within mixed clusters. Genes are categorized as cross-tissue (correlated in both tissues) or tissue-specific (correlated in only one tissue or opposing directions between tissues). b, Number of DE genes identified within tissue-specific clusters, grouped by their predominant tissue of origin (TI or blood). Clusters with >10 DE genes are labeled in (a) and (b). c, Boxplots showing per-sample abundance as a function of the Simple Endoscopic Score for Crohn’s disease at the terminal ileum region (TI-SES-CD) for clusters showing significant tissue-specific associations with mucosal inflammation.

To assess whether changes in immune cell abundance also reflected mucosal inflammation, we analysed the relationship between cluster proportions and TI-SES-CD scores (Methods). Among the mixed clusters, only the abundance of classical monocytes in the blood showed a significant negative association with inflammation (Fig. 3c and Supplementary Table 6), similar to a previous study reporting a negative association between classical monocytes and disease duration in CD^20^. In the ileum, the frequency of *SOD2^+^* intermediate monocytes and plasmablasts was positively associated with inflammation. For tissue-specific clusters, only those from the TI, specifically the four plasma cell clusters and *ITGA4^+^* macrophages, showed positive associations linked with inflammation. These results support the conclusion that transcriptional and compositional changes in response to inflammation are restricted to the local tissue compartment, with the exception of ex-Trms and *SOD2^+^* intermediate monocytes.

### Mucosal inflammation correlates with cytokine signalling in Trms and ex-Trms

To investigate the functional relevance of the cross-tissue inflammation-associated genes identified in Trms, we performed gene set enrichment analysis (GSEA) (Methods). In CD4⁺ Trms, genes upregulated with increasing inflammation were enriched for interferon gamma (IFNγ) signalling (adjusted p = 1×10^-3^), while the pro-inflammatory *KLRB1^+^CCR5^+^* subset showed enrichment in TNFR2 (adjusted p = 1×10^-2^) and interleukin signalling pathways (adjusted p = 2×10^-2^) (Fig. 4a and Supplementary Table 7). These findings suggest that Trms respond to inflammation through coordinated activation of cytokine-driven programs, with distinct emphases across subsets.

Several cytokine-related genes linked to inflammation in Trms, including *IL12RB2, IL1R1, IL21R, JAK3, IL7, SOCS3, STAT1, TNFRSF18, TNFRSF25*, and *TNFRSF4*, were also upregulated in other tissue-resident immune populations such as *ITGA4⁺* macrophages and Trm Tregs (Fig. 4a–b). Both cell types play established roles in IBD pathogenesis^21–23^, and this transcriptional overlap points to a broader, inflammation-associated cytokine network spanning multiple immune lineages in the intestinal mucosa.

**Fig. 4.**
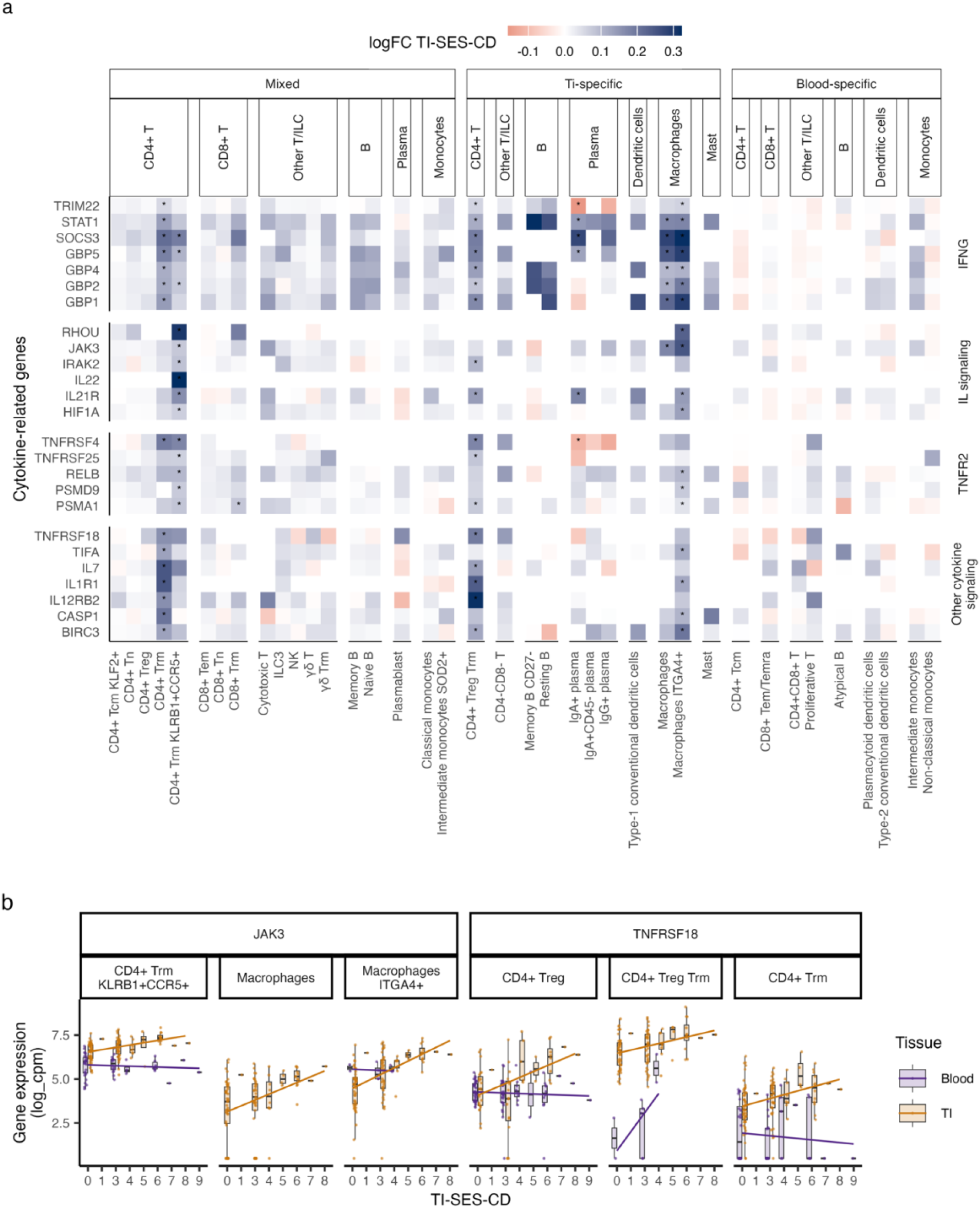
Differential cytokine expression is associated with mucosal inflammation in immune cells. a, Heatmap displaying cytokine-related genes identified through the REACTOME pathway analysis of inflammation-associated genes across tissue-resident memory (Trm) clusters. Colors represent the log fold-change (logFC) correlation with the Simple Endoscopic Score for Crohn Disease at the terminal ileum region (TI-SES-CD). For mixed clusters, the logFC values were calculated across both tissues; while for tissue-specific clusters, only cells from the predominant tissue were included. Asterisks indicate significant correlations (FDR<0.05). b, Boxplots showing the expression levels of *JAK3* and *TNFRSF18* across Trm T-cell subsets and other significantly correlated clusters, colored by tissue origin (TI or blood).

Notably, despite their presence in the circulation, ex-Trms retained these cytokine response signatures, mirroring their tissue counterparts. This preservation of mucosal inflammation-linked transcriptional programmes reinforces their potential utility as a rare but informative blood-based proxy for gut inflammation.

### IFNγ signalling and Treg migration correlate with inflammation in ileal cells

As blood-specific clusters lacked any inflammation-associated genes, we next focused on tissue-specific clusters from the TI. Among 382 genes that significantly correlated with TI-SES-CD, 21 were restricted to a single immune lineage (e.g., T/ILC cells, B cells, or myeloid), 14 were shared across lineages and the remainder were cluster-specific (Supplementary Fig. 5a), highlighting both shared and cell-type-specific immune responses to mucosal inflammation.

Several genes showed inflammation-associated expression in T cells. For example, *PDE4D* was positively correlated with TI-SES-CD scores in central memory CD4⁺ T cells (Tcm) with high *KLF2* expression and effector memory CD8⁺ T cells (Tem) (Supplementary Fig. 5b). In contrast, *PDE4B* was associated with inflammation across lineages, showing positive correlation in Tregs and naive B cells (Supplementary Fig. 5c). While both PDE4-related genes have been linked to endoscopic disease severity (the Mayo score) of ulcerative colitis (UC) via bulk RNA-sequencing of colonic biopsies^24^, our single-cell data extend these findings to CD, indicating T and B lymphocytes as the likely cell types driving these associations. Both genes encode isoforms of phosphodiesterase-4 (PDE4), an enzyme that regulates intracellular cyclic AMP (cAMP). By modulating cAMP degradation, PDE4 influences cytokine production (e.g., IL-2) after TCR engagement. PDE4 inhibitors are approved to treat atopic dermatitis and psoriasis^25^ and are in clinical trials for IBD^26^.

Another cross-lineage gene of interest is *SLAMF1 (CD150)*, which showed positive association with TI-SES-CD scores in mucosal Tregs and memory B cells (Supplementary Fig. 5d). In mice, *SLAMF1* knockout reduces the development of colitis^27^, suggesting that *SLAMF1* not only serves as a biomarker of mucosal inflammation in these populations but may also contribute to IBD pathogenesis. Similarly, the identification of *GBP5* as inflammation-associated in specific cell types illustrates how single-cell resolution can reveal context-specific immune signals that are missed in bulk tissue. Previous studies identified *GBP5* expression in colonic tissues as correlated with SES-CD scores but not with the UC Endoscopic Index of Severity, with its expression confined to immune cells where it regulates cytokine and chemokine production^28^. In our single-cell data, we found significant associations in naive B cells and Tregs (Fig. 5a-b). Together, these results underscore the value of our dataset in revealing tissue-specific and cross-lineage genes associated with mucosal inflammation and their cellular associations.

**Fig. 5.**
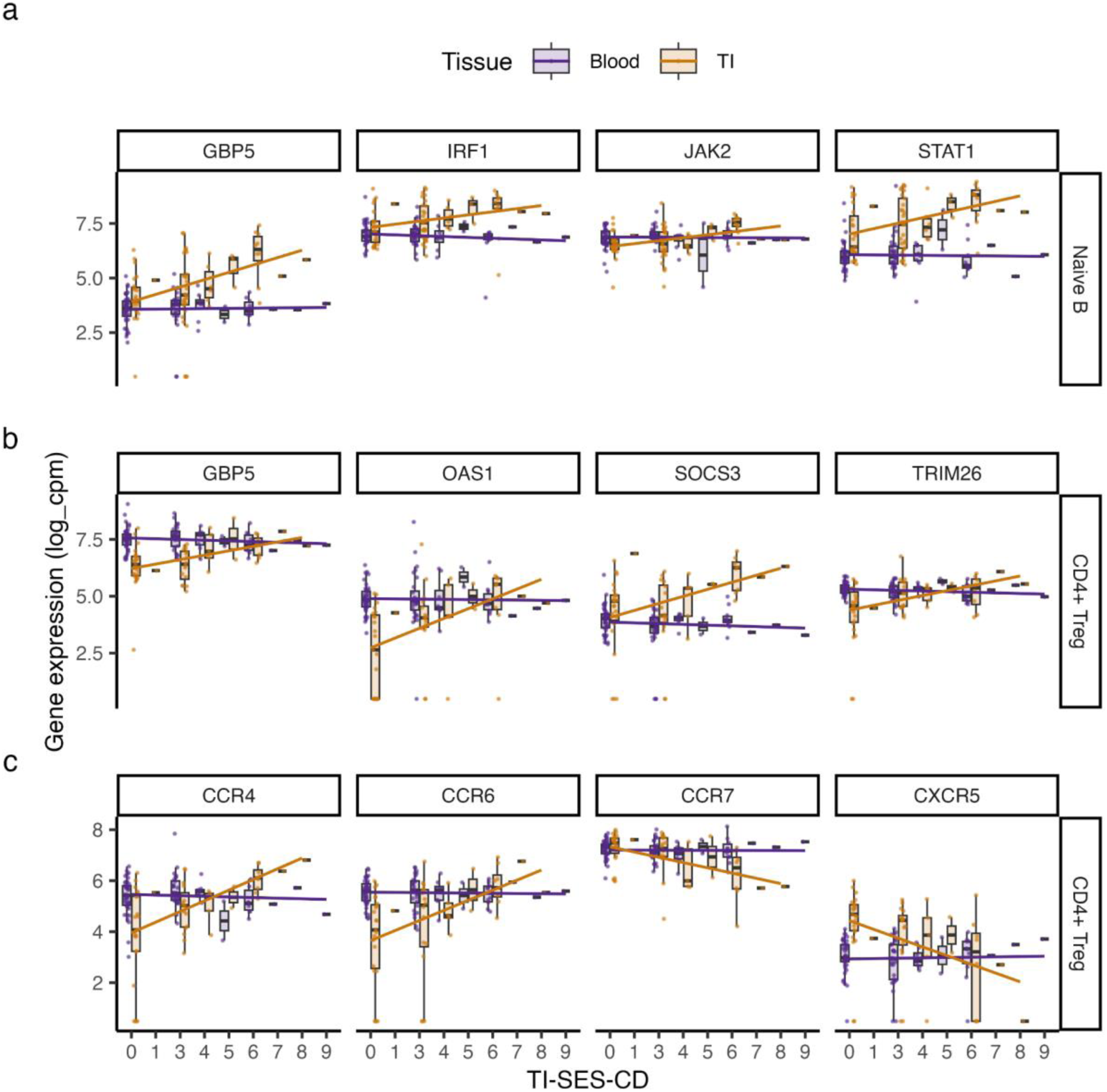
Ileal-specific correlations with mucosal inflammation highlight IFNγ signaling and T cell migration pathways. a-c, Boxplots showing the expression of selected tissue-specific, differentially expressed genes (FDR<0.05) involved in IFNγ signaling and chemokine receptor pathways in naive B cells (a) and Tregs (b-c) from ileal and circulating immune cells. Expression is plotted against each patient’s Simple Endoscopic Score for Crohn Disease at the terminal ileum region (TI-SES-CD).

To assess mucosa-specific pathways associated with inflammation, we performed GSEA for each ileal cluster, ranking genes by the strength of their correlation with TI-SES-CD scores. This revealed significant enrichment of IFNγ signalling in both naive B cells (adjusted p = 1×10^-6^) and Tregs (adjusted p = 2.61×10^-2^) (Supplementary Fig. 5e and Supplementary Table 7), potentially reflecting a shift in their functional state under inflammatory conditions. In naive B cells, IFN-associated genes included *STAT1* and *JAK2* (Fig. 5a), key components of the JAK-STAT pathway targeted by several licensed IBD therapies. These findings suggest that IFNγ-responsive gene programmes are active in naive B cells during inflammation, consistent with prior in vitro evidence linking this pathway to altered B cell differentiation and function^29,30^. In Tregs, IFNγ-related genes included *SOCS3* (Fig. 5b), a negative regulator of cytokine signalling. Overexpression of *SOCS3* has been shown to impair T-cell proliferation and suppressive function and to reduce expression of *FOXP3* and *CTLA-4* in vitro^31^. These findings suggest that IFNγ exposure in the inflamed intestine may impair Treg suppressive function and alter B cell differentiation, potentially amplifying the inflammatory response.

Beyond IFN enrichment, GSEA of inflammation-correlated genes in ileal Tregs revealed enrichment of chemokine receptor pathways (adjusted p = 3×10^-3^) (Supplementary Fig. 5e), suggesting that genes linked to Treg migratory potential are transcriptionally associated with inflammation. Here, *CCR4, CCR6, CXCR6* and *CXCR3* were positively correlated with TI-SES-CD scores, while *CCR7* and *CXCR5*, which are involved in lymph node^32^ and germinal centre homing^33^, showed negative correlations (Fig. 5c). These patterns suggest a shift from central lymphoid homing toward peripheral tissue adaptation in the inflamed intestine. Both chemokine receptor genes and IFNγ-associated genes showed similar correlation patterns in Tregs with a Trm phenotype (Supplementary Fig. 6), indicating a shared transcriptional response to mucosal inflammation.

### Tissue-specific programs are maintained across cell types and inflammation

To investigate how immune cell transcriptomes differ between tissue compartments independently of inflammation, we compared gene expression profiles between ileal and circulating cells within each mixed cluster (Methods). This analysis identified 18,334 differentially expressed (DE) genes between tissues (Fig. 6 and Supplementary Table 8). Among these, 15,158 genes were significant in more than one cluster, and 7,947 genes exhibited opposing directions of effect in at least one cluster (Supplementary Fig. 7a). However, these discordant cases were driven by a small number of clusters, while the majority of cell types demonstrated consistent directions of effect (Supplementary Fig. 7b).

**Fig. 6.**
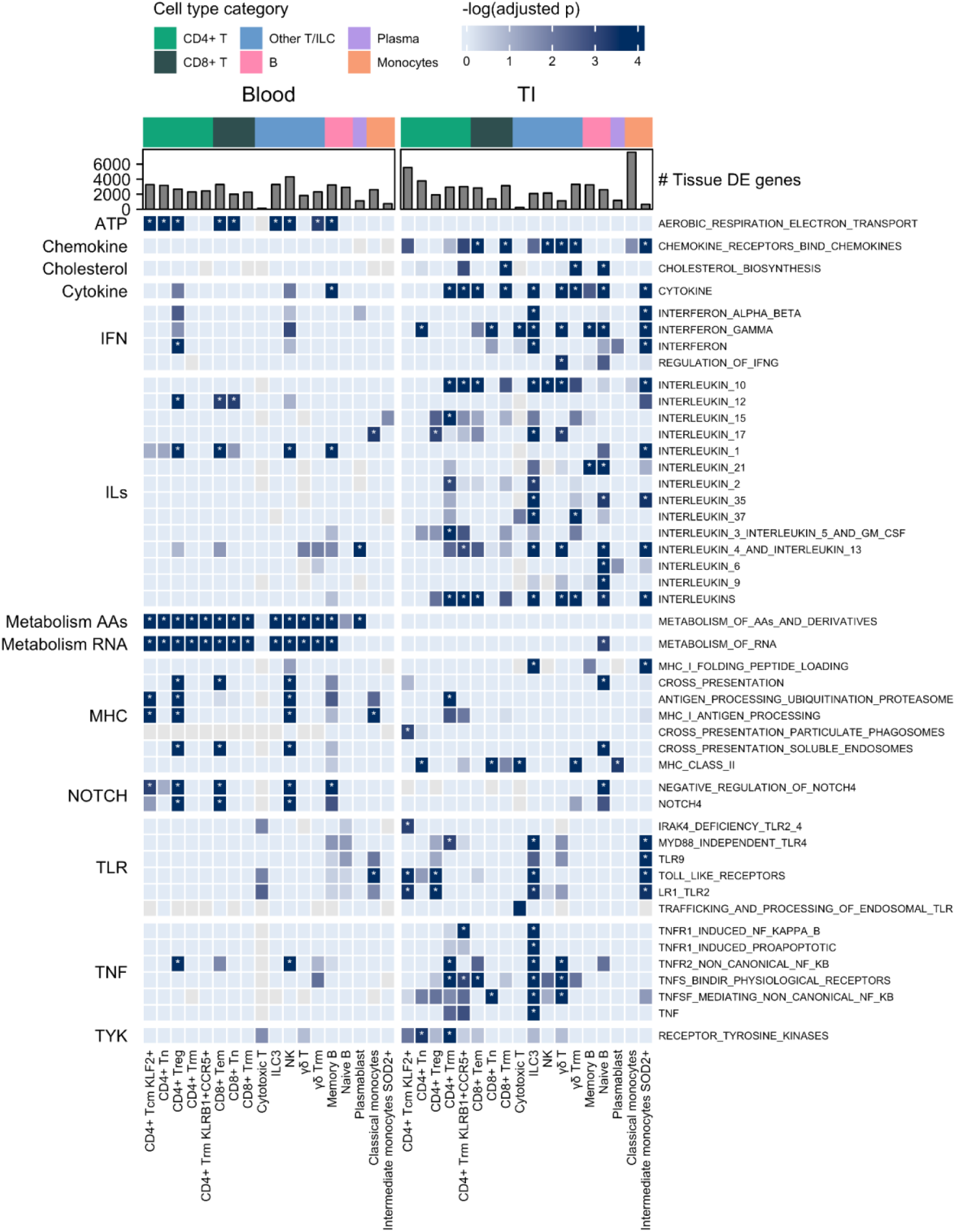
Tissue-specific pathway enrichment in Crohn’s disease. Heatmap showing metabolic and immune-related pathways significantly enriched in tissue-specific differentially expressed (DE) genes, identified through REACTOME pathway analysis. Columns represent the enrichment in immune cell clusters from genes upregulated in circulating (left) and mucosal (right) compartments, grouped by cell type category. Rows indicate significantly enriched pathways, grouped by functional category. Bar plots above each column show the number of DE genes per cluster. Color intensity reflects pathway enrichment significance and asterisks denote statistically significant associations (adjusted p < 0.05). Functional categories include interferons (IFN), interleukins (ILs), metabolism of amino acids (Metabolism AAs) and RNA (Metabolism RNA), major histocompatibility complex (MHC), Notch signaling (NOTCH), Toll-like receptors (TLR), tumor necrosis factor signaling (TNF), and tyrosine kinases (TYK).

To assess whether these differences reflect canonical features of tissue residency, we curated a list of 37 genes previously linked to tissue-resident immune cell identity across diverse tissues and lineages^34^. Of these, 34 genes were DE between circulating and ileal cells, with 20 showing consistent patterns across multiple lineages (Supplementary Fig. 7c). This supports the existence of a conserved tissue-specific transcriptional signature in the TI, even under inflammatory conditions. Among the most consistently DE genes were key Trm markers: *CD69*, *KLF2*, *SELL*, and *ITGA1*, suggesting these serve as global markers of tissue-localized immune cells beyond T cells. In contrast, 14 genes exhibited inconsistent patterns. For example, *S1PR1* and *LEF1* were more highly expressed in circulating lymphoid cells than in their mucosal counterparts, but showed the opposite trend in monocytes. Similarly, *RUNX3* was upregulated in ileal monocytes, T cells, and B cells, yet downregulated in ileal NK cells.

Beyond canonical residency markers, ileal immune cells exhibited distinct metabolic and signalling programmes compared to their circulating counterparts^3^. Circulating T and B cells showed higher expression of genes linked to aerobic respiration, amino acid metabolism, and RNA synthesis, consistent with a quiescent, recirculating phenotype. In contrast, ileal CD8⁺ Trms, γδ Trms and naive B cells upregulated genes associated with cholesterol biosynthesis (Fig. 6 and Supplementary Table 9). This pathway has been previously linked to Th1 and Th17 differentiation^35^ and aligns with elevated IFNγ and IL-17A production in the lamina propria of CD patients^36–38^. In addition to metabolic differences, ileal immune cells displayed higher expression of genes in key IBD-related pathways, including IFN signaling, tumor necrosis factor (TNF) signaling, toll-like receptor (TLR) pathways, receptor tyrosine kinases (RTK), and NOTCH4 signaling. Enrichment of chemokine, cytokine, and interleukin pathways was also observed across multiple lineages. Many of these pathways are targets of existing IBD therapies or represent active areas of drug development, underscoring the translational relevance of tissue-specific immune programs.

### Cytokine regulation is associated with tissue-specific and inflammatory differences

To explore how tissue-specific transcriptional programmes intersect with inflammation severity, we compared DE genes between tissues to those whose expression in the ileum was correlated with TI-SES-CD scores. This analysis identified 266 genes whose expression was both tissue-specific and associated with inflammation extent, suggesting overlap between residency-associated and inflammation-induced transcriptional states.(Supplementary Fig. 7d). GSEA of these genes revealed significant associations with IFNγ signaling in naive B cells (adjusted p = 7×10^-7^) and Tregs (adjusted p = 1×10^-3^), as well as cytokine signaling in B cells (adjusted p = 6×10^-3^) (Supplementary Table 10). These findings indicate that tissue-specific cytokine regulation, particularly IFNγ signaling, is a key feature distinguishing ileal immune cells from their circulating counterparts while also shaping inflammation severity within the ileum.

Notably, several tissue-specific genes did not show significant correlation with TI-SES-CD scores (Supplementary Fig. 7d), suggesting that some transcriptional differences between tissues reflect stable features of immune cell identity. However, these unshared genes may also include inflammation-associated transcripts that were not detected due to limited statistical power. Specifically, tissue-specific DE genes were influenced by the number of aggregated cells and samples within each cluster, whereas the identification of inflammation-associated genes was not (Supplementary Fig. 8). This difference in statistical power highlights that while cytokine-driven transcriptional programs play a major role in both tissue residency and inflammation, additional pathways may contribute but remain undetected in this dataset.

## Discussion

Blood-based profiling offers a minimally invasive route to patient stratification in CD, but how well circulating immune cells reflect the biology of gut inflammation has remained unclear. This question is particularly pressing given the use of transcriptomic and proteomic signatures derived from peripheral blood in biomarker discovery and drug development. To address this, we analysed paired single-cell data from blood and TI, revealing that, aside from a rare population of circulating ex-Trms, inflammation-associated transcriptional signatures were largely confined to the gut. Similar findings were reported in a single-cell study of 29 patients with immune checkpoint inhibitor-associated colitis, where colitis-related immune signatures were present in intestinal tissue but absent from blood^39^. This convergence across distinct inflammatory conditions underscores the limited capacity of circulating immune cells to capture local inflammation. Our larger CD cohort (n = 125) extends these findings and reinforces the tissue-restricted nature of inflammatory responses.

More importantly, the discovery that a subset of circulating T cells retains transcriptional features typically restricted to Trms, sheds new light into immune surveillance in CD. While murine studies have documented egress of gut-derived Trms into circulation, analogous populations in humans have thus far only been described in the skin^16,17^. Our data provides evidence for a similar phenomenon in the human gut: circulating CD4⁺ and CD8⁺ T cells expressing canonical Trm markers, including *ITGAE*, *ITGA1*, *CD101*, and the gut-homing receptor *CCR9*. These cells were transcriptionally similar to ileal Trms and were the only circulating population whose gene expression profiles correlated with mucosal inflammation, including enrichment for IFNγ, TNFR2, and interleukin signalling pathways. Importantly, we identified comparable ex-Trm-like cells in independent blood single-cell datasets, suggesting this phenomenon may generalise beyond CD. While their precise tissue of origin cannot be definitively established, their expression patterns and inflammation-associated signatures are consistent with prior gut residency. These findings position ex-Trms as a rare but informative blood-based correlate of mucosal inflammation.

Interestingly, a prior study of CD4⁺ T cells in juvenile idiopathic arthritis found that while most circulating cells were transcriptionally distinct from their synovial counterparts, a small subset correlated with disease activity and showed substantial clonal overlap with synovial T cells^40^. Although Trm markers were not assessed, this population may represent an ex-Trm-like subset. Alongside our detection of CD8⁺ ex-Trms in independent blood single-cell datasets from other inflammatory contexts, these findings raise the possibility that ex-Trms may serve as broadly applicable biomarkers of chronic tissue inflammation. This could be particularly relevant for diseases where tissue sampling is not routinely feasible. However, their low abundance may limit their predictive value in clinical settings and complicate efforts to develop scalable diagnostic assays. Nonetheless, ex-Trms may offer a rare but functionally informative snapshot of mucosal T cell biology, enabling mechanistic studies without direct tissue sampling.

While circulating cells largely failed to capture inflammation-associated transcriptional changes, mucosal immune populations revealed robust and cell type-specific responses to local inflammatory cues. In particular, Tregs and naive B cells demonstrated strong associations with IFNγ-responsive gene programmes, including core JAK–STAT signalling components such as *STAT1* and *JAK2*. These findings suggest that cytokine exposure in the intestinal microenvironment induces lineage-specific transcriptional reprogramming, with potential consequences for both regulatory and effector cell function. For example, in Tregs, expression of *SOCS3*, a negative regulator of cytokine signalling known to impair FOXP3 stability and suppressive capacity^31^, was tightly linked to inflammation severity, pointing to functional destabilisation of the regulatory compartment. Enrichment of chemokine receptor pathways in these same cells further supports the emergence of a migratory, inflammation-adapted Treg state in the inflamed gut. Together, these findings provide a high-resolution view of how chronic inflammation remodels mucosal immune cell states and reinforce the central role of cytokine-driven transcriptional networks in shaping tissue-level immune phenotypes in CD. By resolving these programmes at single-cell resolution, our study clarifies how distinct immune lineages respond transcriptionally to a shared inflammatory environment, an insight that bulk profiling cannot capture.

While this study provides a comprehensive atlas of mucosal and circulating immune cells in CD, several limitations should be acknowledged. First, although the cohort size is substantial compared with other scRNAseq datasets, the relatively low abundance of some cell states may have limited our power to detect more subtle associations with inflammation. Second, while SES-CD scores have high interobserver agreement and correlate to clinical symptoms and serum C-reactive protein levels^41^, other clinically relevant dimensions of CD such as extra-intestinal manifestations, symptom burden, or treatment response may not be captured by SES-CD. Additionally, the cross-sectional nature of our sampling prevents assessment of immune state transitions over time. Future work incorporating longitudinal sampling, functional assays, and integration with additional molecular layers (e.g. proteomics or microbiome) will be important to further resolve the immune mechanisms underlying disease progression, treatment response, and inter-patient heterogeneity.

Much of the existing work on tissue-specific immune differences has focused primarily on T cells and has been limited to healthy individuals^2–4,11^. By contrast, our study presents a high-resolution single-cell resource from a large cohort of CD patients, enabling the first systematic comparison of transcriptional and compositional immune phenotypes across tissues and inflammation states under chronic inflammation. The matched sampling design allowed us to disentangle local and systemic influences on immune cell states, revealing that transcriptional signatures of inflammation are overwhelmingly tissue-restricted, with the exception of ex-Trms. These findings build on and extend prior work by clarifying which immune features are shared between compartments, which are tissue-specific, and which track with inflammation severity. More broadly, they reinforce the importance of studying immune variation in disease-relevant tissues and provide a framework for future efforts to identify biomarkers and therapeutic targets in IBD.

## Data Availability

The raw sequencing data, processed data, and code used for analysis will be publicly available upon publication.

## Acknowledgments

This research was supported by the NIHR Cambridge Biomedical Research Centre (BRC-1215-20014). The views expressed are those of the authors and not necessarily those of the NIHR or the Department of Health and Social Care. This research was funded in part by the Wellcome Trust [Grant numbers 206194 and 108413/A/15/D] and The Crohn’s Colitis Foundation Genetics Initiative [Grant numbers 612986 and 997266]. We thank the Human Genetics Informatics team for their support in the project and all individuals who kindly donated samples and their time to the study.

## Author contributions

Statistical analysis and manuscript drafting, L.R.N; Sample collection, W.G., B.B., and C.Q.C.; Sample processing, M.S., M.G., J.O., N.W., M.X.H., J.S.,R.E.M., and C.C.M; Critical discussion, G.R.J., B.T.H., T.A., M.P., T.R. and C.A.A; Data processing T.A.,S.L., and V.I; Writing – Review & Editing, L.R.N, G.R.J, B.T.H.,C.C.M, T.R. and C.A.A.; Conceptualization and Supervision, T.R., C.A.A.

## Declaration of interests

B.T.H. has received speaker fees from BridgeBio. C.A.A. has received research grants or consultancy/speaker fees from Genomics plc, BridgeBio, GSK and AstraZeneca. T.R. has received research/educational grants and/or speaker/consultation fees from Abbvie, Arena, Aslan, AstraZeneca, Boehringer-Ingelheim, BMS, Celgene, Ferring, Galapagos, Gilead, GSK, Heptares, LabGenius, Janssen, Mylan, MSD, Novartis, Pfizer, Sandoz, Takeda and UCB. R.M. is an employee at Relation Therapeutics.

## Methods

### Sample collection

This study was approved by the National Health Service (NHS) Research Ethics Committee (Cambridge South, REC ID 17/EE/0338). Written informed consent was obtained from all participants.

A total of 125 CD patients who were not taking probiotics or antibiotics were recruited at Addenbrooke’s Hospital, Cambridge, UK. For 70% (N = 88) of patients, TI pinch biopsies and blood samples were collected simultaneously during endoscopic assessment, while 7 patients provided only blood and 30 patients only TI.

Mucosal inflammation was quantified using the TI-SES-CD score, which sums three inflammatory subscores of the SES-CD score: ulcer size, ulcerated surface area and inflamed surface area, excluding stenosis. The scores were adjudicated by a single blinded central reader and each subscore ranges from 0 (no inflammation) to 3, with a maximum possible score of 9 indicating severe inflammation in the TI (Supplementary Table 1).

TI pinch biopsies were collected from the participants and immediately placed into pre-chilled Hanks Balanced Salt Solution (HBSS) without Mg^2+^, Ca^2+^, and phenol red. Samples were kept on ice and transported to the Sanger Institute for further processing. Peripheral blood was collected in K2 EDTA tubes, also kept on ice, and transferred alongside the TI samples, typically within one hour of collection.

### Single-cell RNA isolation and sequencing

TI biopsies were dissociated and processed into single cell suspension following the protocol described in Krzak et al^9^. Briefly, samples were incubated on ice with BLP protease/2 mM EDTA and two apoptotic inhibitors (Rho Kinase and Caspase inhibitors) for 30 minutes, followed by enzymatic digestion with collagenase at room temperature for 10 minutes. In parallel, peripheral mononuclear blood cells (PBMCs) were isolated from 1-2 ml of blood using the EasySep Direct Human PBMC Isolation Kit (Cat. 19654 StemCell Technologies) following manufacturer’s protocol. For both tissues, 9,000 cells were loaded into 10x Chips lane to capture 6000 cells for scRNA-seq using 10X Genomics Next GEM single cell 3’ kits (v3.0 and v3.1), following the manufacturer’s protocol.

### Single-cell RNA-seq processing and quality control

FASTQ reads were demultiplexed and aligned to the human reference genome (GRCh38-3.0.0 reference file distributed by 10X Genomics) using Cell Ranger (v7.2.0), generating a unique molecular identifier (UMI) count matrix of cells by genes. To correct for ambient RNA contamination and identify droplets, CellBender (v2.1)^42^ was applied and potential multiples were removed using Scrublet (v0.2.1)^43^. Subsequent processing and management of the single cell data was performed using the scanpy library (v1.9.8)^44^. For both tissues, cells were filtered by selecting those with a percentage of reads mapping to the mitochondrial genome below 17% and with 400 to 8000 genes expressed at ≥ 1 count. Genes expressed in less than 5 cells were also removed.

### Selection of variable genes, dimensionality reduction, clustering and annotation

To leverage information from existing single-cell atlases, we predicted cell type labels using five published atlases: a PBMC atlas of healthy individuals^45^, an atlas of immune populations across 20 tissues^2^ and three gut-related atlases from healthy and IBD patients^9,46,47^. The original cell type models created by the authors were used and cell type information was harmonized across these five models into nine broad immune categories: Plasma B cells, B cells, T/ILC cells, Macrophages, Monocytes, Dendritic cells, Mast cells, other immune cells and non-immune cells.

Since TI samples contained non-immune cells, we applied a voting-based approach inspired by work from Ergen et al^48^ to filter them out. Cell type labels for TI samples were predicted using the three gut atlases, and the most commonly assigned broad category was selected. If no consensus was reached, the label with the highest confidence score was assigned. To confirm the immune categories, we visualized marker gene expression from an independent atlas of circulating immune cells across conditions^49^ and canonical epithelial markers *(CDH1, KRT19, EPCAM)* (Supplementary Fig. 9). Cells identified as non-immune were excluded from further analysis and immune cells from both tissues were combined. Then, gene expression was normalized by the total counts, followed by log-transformation and the 5,000 most highly variable genes across samples and tissues were selected using the flavour of seurat and sample ID as batch key.

Cell type annotation was performed using a recursive top-down approach. First, we integrated cells from both tissues using scVI (v1.1.5) with sample ID as the batch key and applied the recommended non-default parameters from their tutorial (n_latent= 30, n_layers=2, gene_likelihood=’nb’). After computing neighbors using the scVI embedding, cells were clustered at a low resolution (resolution = 0.01) to define major immune lineages using the cell type predictions and immune marker expression. In total, five clusters were identified: one T-cell based, two B-cell clusters, a myeloid cluster and another cluster with no concordant annotation (Supplementary Fig. 10a-b). While all the other clusters showed agreement across all the five methods, this one was annotated as Platelets by the PBMC reference, Mast cells by the cross-immune atlas and two gut-atlases while the other gut-atlas annotated it primarily as T cells (Supplementary Table 2). Consistent with this, marker gene identification (sc.tl.rank_genes_groups(method=’t-test’)) showed expression of myeloid *(RUNX1, GATA2, CSF2RB, JAK2, TMEM154)*, lymphoid *(CAMK1D, CHST11, GNAQ)* and Mast and platelet-related genes *(GAB2, TBXAS1, GATA2)*. 97% of these cells were circulating (Supplementary Fig. 10c) and the cluster had expression of *CD45* and no expression of epithelial gene markers, suggesting these cells were immune (Supplementary Fig. 10b). Compared to the other clusters, this was a small cluster representing only 6,749 out of 886,979 cells (0.7%) and less than 50 cells per sample, meaning that further sub-clustering of this population was impractical. Additionally, this cluster had significantly less number of expressed genes and total counts compared to the other clusters (Supplementary Fig. 10d), suggesting these are low quality cells which were further excluded in the analysis.

The other three immune lineages (T/ILC cells, B cells and myeloid) underwent a second round of integration with scVI, followed by clustering to define finer cell states. As previously described^9^, the optimal number of clusters for each lineage was determined in a data-driven manner. Each lineage was clustered at resolutions ranging from 0.3 to 1 (in 0.1 intervals), and a single layer neural network was trained on two-thirds of the data to predict cluster labels in the remaining third. The final clustering resolution was selected based on Matthews correlation coefficient (MCC) ≥0.75. The chosen resolutions for the final cell clusters were: 0.7 for B cells, 0.7 for T/ILC cells and 0.4 for myeloids cells.

For annotation, we considered the cell type abundance at both the cell and sample level and the two most represented predicted cell type labels per method and verified them using canonical immune markers and additional literature-based marker genes (Supplementary Fig. 2 and Supplementary Table 2). For T cells, we also integrated predictions from a meta-analysis of T cell subsets across 38 tissues and five diverse disease contexts^50^. Low quality clusters based on the number of cells or expression of marker genes from other lineages were also removed.

### Validation of ex-Trms across other blood single-cell datasets

Using the blood-derived data and our cell type annotations, a CellTypist (v1.6.2)^2,18^ model with the normalized gene expression levels across all genes was trained. Next, the raw gene expression data from the CD8+ T cell single-cell meta-analysis was obtained via their Zenodo repository (v3) (https://zenodo.org/records/13382785)^19^. The dataset was subsetted to PBMC-derived cells using the meta_tissue_type column. Genes were normalized, and cell type predictions were generated using our CellTypist model. High confident predictions were determined with confidence > 0.5.

### Differential gene expression analysis

To investigate gene expression patterns of immune cells in relation to both mucosal inflammation (TI-SES-CD scores) and tissue differences (blood vs TI), we performed a pseudobulk differential gene expression (DGE) analysis using the dream model from the variancePartition package (v1.32.5)^51^. This approach uses a precision-weighted linear mixed model to account for repeated measurements within the same patient, while improving statistical power.

### Pseudobulk processing and normalization

Gene expression was aggregated by summing counts across all cells from the same patient, tissue, and cluster using the adpbulk library (https://github.com/noamteyssier/adpbulk). Given that DGE power is associated with sample size^52^, only clusters detected in at least 40 patients and samples with ≥10 cells per cluster were considered. To ensure balanced comparisons in the DGE analysis, clusters were classified as mixed (present in both blood and TI in ≥25% of samples per tissue) or tissue-specific (predominantly found in one tissue with >75% from a single tissue). This classification was performed at the sample level to avoid biases introduced due to experimental constraints such as differences in sample processing and the presence of non-immune cells in the TI. The thresholds selected were informed by the distribution of tissue composition per cluster, including the locations of the local maxima and minima in the smoothed density curve, which reflected the boundaries between tissue-specific and mixed clusters (Supplementary Fig. 11).

For each cluster, lowly expressed genes were filtered using edgeR (v4.0.16) (filterByExpr(min.count = 5, min.prop = 0.4, min.total.count = 15)), incorporating the relevant covariate of interest. For mixed clusters, tissue was included as an additional covariate.

### Genes associated with mucosal inflammation (TI-SES-CD scores)

To identify genes correlated with mucosal inflammation, we performed DGE analysis within each cluster. In mixed clusters, two sets of genes were identified:

1. Cross-tissue associations: genes correlated with TI-SES-CD scores in both blood and TI, with the same direction of effect. This was assessed by modeling TI-SES-CD scores while adjusting for the tissue of origin.
2. Tissue-specific associations: genes where the correlation with TI-SES-CD scores differed between blood and TI. These genes were identified using an interaction term between TI-SES-CD scores and tissue. To systematically characterize the direction of effect for the significant genes, a DGE analysis was performed separately for each tissue. A gene could be significant in both tissues with opposing directions of effect or significant in only one tissue.

In tissue-specific clusters, we only tested for an association with TI-SES-CD scores within the predominant tissue.

### Genes with tissue differences (blood vs TI)

For mixed clusters, we identified genes where the expression levels differed between circulating and mucosal immune cells within each cluster, while accounting for the degree of inflammation.

### General statistical framework

For each cluster and DGE analysis, a voom-style normalization for pseudobulk counts was applied and a regression model was fitted to test differential expression with the covariate of interest. All models included covariates to adjust for biological (sex, age, and medication status) and technical (mitochondrial gene percentage and number of aggregated cells per sample) sources of variation. To account for repeated measures, patient ID was included as a random effect in mixed clusters, with tissue modelled as fixed effects. Empirical Bayes shrinkage was applied to the linear mixed models, and multiple testing correction was performed using the Benjamini–Hochberg method. Significant genes were defined as FDR<0.05.

### Differential abundance analysis

To assess whether immune cell proportions are associated with mucosal inflammation, we performed a differential abundance (DA) analysis. Donors with less than 500 total immune cells were excluded and for each patient, the number of cells per tissue and cluster were calculated.

Following the same statistical approach as our DGE analysis, we stratified the analysis based on the tissue composition of each cluster and applied separate models. Sex, age and medication status were included as covariates and for mixed clusters, patient ID was modelled as random effect to account for repeated measures, while tissue was included as a fixed effect.

For mixed clusters, we identified two types of associations with mucosal inflammation:

1. Cross-tissue associations: clusters where abundance was correlated with TI-SES-CD scores in both blood and TI, with the same direction of effect. These were identified by modeling the TI-SES-CD scores while adjusting for tissue identity.
2. Tissue-specific associations: clusters where the relationship with TI-SES-CD scores differed between blood and TI, identified using an interaction term between the TI-SES-CD scores and tissue.

For tissue-specific clusters, associations with TI-SES-CD scores were tested only within the predominant tissue.

Multiple testing correction was applied across all tested clusters, and significant associations were defined as adjusted p-value < 0.05.

### Gene set enrichment analysis

Gene set enrichment analysis was performed using gProfiler2 (v0.2.3)^53^ to identify Reactome pathways enriched among the differentially expressed genes. Curated Reactome pathways were downloaded from the Human MSigDB v2024 collection^54^. Genes were divided by their logFC direction and ordered based on their effect size (logFC) to generate GSEA style p-values. In the case of significant genes derived from the interaction analysis (tissue-specific associations with TI-SES-CD scores), adjusted p-values were used for ordering. Significant terms were selected with an adjusted p-value < 0.05 and more than 3 significant genes in the pathway. For background, genes that were tested in the DGE analysis were used.

## Notes

### Author Declarations

The Cambridge South Research Ethics Committee of the National Health Service (REC ID 17/EE/0338) gave ethical approval to this work.

## References

1. de Souza, H. S. P. & Fiocchi, C. Immunopathogenesis of IBD: current state of the art. Nature Reviews Gastroenterology & Hepatology 13, 13–27 (2015).

2. Domínguez Conde, C., et al. Cross-tissue immune cell analysis reveals tissue-specific features in humans. Science 376, eabl5197 (2022).

3. Wells, S. B., et al. Multimodal profiling reveals tissue-directed signatures of human immune cells altered with age. bioRxiv (2024) doi:10.1101/2024.01.03.573877.

4. Poon, M. M. L. et al. Tissue adaptation and clonal segregation of human memory T cells in barrier sites. Nat Immunol 24, 309–319 (2023).

5. Boland, B. S., et al. Heterogeneity and clonal relationships of adaptive immune cells in ulcerative colitis revealed by single-cell analyses. Sci Immunol 5, (2020).

6. Chiarolla, C. M. et al. Pro-inflammatory NK-like T cells are expanded in the blood and inflamed intestine in Crohn’s disease. Mucosal Immunol 18, 162–175 (2025).

7. Globig, A.-M. et al. High-dimensional profiling reveals Tc17 cell enrichment in active Crohn’s disease and identifies a potentially targetable signature. Nat Commun 13, 3688 (2022).

8. Raine, T., Liu, J. Z., Anderson, C. A., Parkes, M. & Kaser, A. Generation of primary human intestinal T cell transcriptomes reveals differential expression at genetic risk loci for immune-mediated disease. Gut 64, 250–259 (2015).

9. Krzak, M. et al. Single-Cell RNA Sequencing of Terminal Ileal Biopsies Identifies Signatures of Crohn’s Disease Pathogenesis. medRxiv 2023.09.06.23295056 (2024) doi:10.1101/2023.09.06.23295056.

10. Pararasa, C. et al. Reduced CD27IgD B Cells in Blood and Raised CD27IgD B Cells in Gut-Associated Lymphoid Tissue in Inflammatory Bowel Disease. Front Immunol 10, 361 (2019).

11. Gray, J. I., et al. Human γδ T cells in diverse tissues exhibit site-specific maturation dynamics across the life span. Sci Immunol 9, eadn3954 (2024).

12. Yokoi, T. et al. Identification of a unique subset of tissue-resident memory CD4+ T cells in Crohn’s disease. Proceedings of the National Academy of Sciences 120, e2204269120 (2023).

13. Behr, F. M. et al. Tissue-resident memory CD8+ T cells shape local and systemic secondary T cell responses. Nature Immunology 21, 1070–1081 (2020).

14. Fonseca, R. et al. Developmental plasticity allows outside-in immune responses by resident memory T cells. Nat Immunol 21, 412–421 (2020).

15. Wijeyesinghe, S. et al. Expansible residence decentralizes immune homeostasis. Nature 592, 457–462 (2021).

16. Klicznik, M. M. et al. Human CD4+CD103+ cutaneous resident memory T cells are found in the circulation of healthy individuals. Science Immunology (2019) doi:10.1126/sciimmunol.aav8995.

17. Strobl, J. et al. Human resident memory T cells exit the skin and mediate systemic Th2-driven inflammation. J Exp Med 218, (2021).

18. Xu, C. et al. Automatic cell-type harmonization and integration across Human Cell Atlas datasets. Cell 186, 5876–5891.e20 (2023).

19. Xue, Z. et al. Integrative mapping of human CD8+ T cells in inflammation and cancer. Nature Methods 1–11 (2024).

20. Kosoy, R. et al. Deep Analysis of the Peripheral Immune System in IBD Reveals New Insight in Disease Subtyping and Response to Monotherapy or Combination Therapy. Cell Mol Gastroenterol Hepatol 12, 599–632 (2021).

21. Na, Y. R., Stakenborg, M., Seok, S. H. & Matteoli, G. Macrophages in intestinal inflammation and resolution: a potential therapeutic target in IBD. Nat Rev Gastroenterol Hepatol 16, 531–543 (2019).

22. Hegarty, L. M., Jones, G.-R. & Bain, C. C. Macrophages in intestinal homeostasis and inflammatory bowel disease. Nature Reviews Gastroenterology & Hepatology 20, 538–553 (2023).

23. Yamada, A. et al. Role of regulatory T cell in the pathogenesis of inflammatory bowel disease. World J Gastroenterol 22, 2195–2205 (2016).

24. Li, H. et al. Targeting PDE4 as a promising therapeutic strategy in chronic ulcerative colitis through modulating mucosal homeostasis. Acta Pharmaceutica Sinica B 12, 228–245 (2022).

25. Baillie, G. S., Tejeda, G. S. & Kelly, M. P. Therapeutic targeting of 3’,5’-cyclic nucleotide phosphodiesterases: inhibition and beyond. Nat Rev Drug Discov 18, 770–796 (2019).

26. Liu, J., Di, B. & Xu, L.-L. Recent advances in the treatment of IBD: Targets, mechanisms and related therapies. Cytokine Growth Factor Rev 71-72, 1–12 (2023).

27. van Driel, B. et al. Signaling lymphocyte activation molecule regulates development of colitis in mice. Gastroenterology 143, 1544–1554.e7 (2012).

28. Li, Y. et al. The Proinflammatory Role of Guanylate-Binding Protein 5 in Inflammatory Bowel Diseases. Front Microbiol 13, 926915 (2022).

29. Frede, N. et al. JAK inhibitors differentially modulate B cell activation, maturation and function: A comparative analysis of five JAK inhibitors in an in-vitro B cell differentiation model and in patients with rheumatoid arthritis. Frontiers in Immunology 14, 1087986 (2023).

30. Rizzi, M. et al. Impact of tofacitinib treatment on human B-cells in vitro and in vivo. Journal of Autoimmunity 77, 55–66 (2017).

31. Pillemer, B. B. L., Xu, H., Oriss, T. B., Qi, Z. & Ray, A. Deficient SOCS3 expression in CD4+CD25+FoxP3+ regulatory T cells and SOCS3-mediated suppression of Treg function. Eur J Immunol 37, 2082–2089 (2007).

32. Menning, A. et al. Distinctive role of CCR7 in migration and functional activity of naive- and effector/memory-like Treg subsets. European Journal of Immunology 37, 1575–1583 (2007).

33. Vanderleyden, I. et al. Follicular Regulatory T Cells Can Access the Germinal Center Independently of CXCR5. Cell Rep 30, 611–619.e4 (2020).

34. Gray, J. I. & Farber, D. L. Tissue-Resident Immune Cells in Humans. Annu Rev Immunol 40, 195–220 (2022).

35. Perucha, E. et al. The cholesterol biosynthesis pathway regulates IL-10 expression in human Th1 cells. Nature Communications 10, 1–13 (2019).

36. Jaeger, N. et al. Single-cell analyses of Crohn’s disease tissues reveal intestinal intraepithelial T cells heterogeneity and altered subset distributions. Nat Commun 12, 1921 (2021).

37. Martin, J. C. et al. Single-Cell Analysis of Crohn’s Disease Lesions Identifies a Pathogenic Cellular Module Associated with Resistance to Anti-TNF Therapy. Cell 178, 1493–1508.e20 (2019).

38. Fuss, I. J. et al. Disparate CD4+ lamina propria (LP) lymphokine secretion profiles in inflammatory bowel disease. Crohn’s disease LP cells manifest increased secretion of IFN-gamma, whereas ulcerative colitis LP cells manifest increased secretion of IL-5. J Immunol 157, 1261–1270 (1996).

39. Thomas, M. F. et al. Single-cell transcriptomic analyses reveal distinct immune cell contributions to epithelial barrier dysfunction in checkpoint inhibitor colitis. Nat Med 30, 1349–1362 (2024).

40. Spreafico, R. et al. A circulating reservoir of pathogenic-like CD4+ T cells shares a genetic and phenotypic signature with the inflamed synovial micro-environment. Ann Rheum Dis 75, 459–465 (2016).

41. Daperno, M. et al. Development and validation of a new, simplified endoscopic activity score for Crohn’s disease: the SES-CD. Gastrointest Endosc 60, 505–512 (2004).

42. Fleming, S. J. et al. Unsupervised removal of systematic background noise from droplet-based single-cell experiments using CellBender. Nat. Methods 20, 1323–1335 (2023).

43. Wolock, S. L., Lopez, R. & Klein, A. M. Scrublet: Computational Identification of Cell Doublets in Single-Cell Transcriptomic Data. Cell Syst 8, 281–291.e9 (2019).

44. Wolf, F. A., Angerer, P. & Theis, F. J. SCANPY: Large-scale single-cell gene expression data analysis. Genome Biol. 19, 15 (2018).

45. Hao, Y. et al. Integrated analysis of multimodal single-cell data. Cell 184, 3573–3587.e29 (2021).

46. Elmentaite, R. et al. Cells of the human intestinal tract mapped across space and time. Nature 597, 250– 255 (2021).

47. Oliver, A. J. et al. Single Cell Integration Characterises Gastric Cell Metaplasia in Inflammatory Intestinal Diseases. bioRxiv 2024.10.11.614415 (2024) doi:10.1101/2024.10.11.614415.

48. Ergen, C. et al. Consensus prediction of cell type labels in single-cell data with popV. Nat Genet 56, 2731–2738 (2024).

49. Jiménez-Gracia, L. et al. Interpretable Inflammation Landscape of Circulating Immune cells. bioRxiv 2023.11.28.568839 (2023) doi:10.1101/2023.11.28.568839.

50. Kotliar, D. et al. Reproducible single cell annotation of programs underlying T-cell subsets, activation states, and functions. bioRxiv 2024.05.03.592310 (2024) doi:10.1101/2024.05.03.592310.

51. Hoffman, G. E. & Roussos, P. Dream: powerful differential expression analysis for repeated measures designs. Bioinformatics 37, 192–201 (2020).

52. Crowell, H. L. et al. muscat detects subpopulation-specific state transitions from multi-sample multi-condition single-cell transcriptomics data. Nature Communications 11, 1–12 (2020).

53. Kolberg, L., Raudvere, U., Kuzmin, I., Vilo, J. & Peterson, H. gprofiler2 -- an R package for gene list functional enrichment analysis and namespace conversion toolset g:Profiler. F1000Research 9, 709 (2020).

54. Subramanian, A. et al. Gene set enrichment analysis: a knowledge-based approach for interpreting genome-wide expression profiles. Proc Natl Acad Sci U S A 102, 15545–15550 (2005).

