## Supplementary figures and supplementary table legends for "Tissue-specific and circulatory immune signatures of mucosal inflammation in Crohn’s disease"

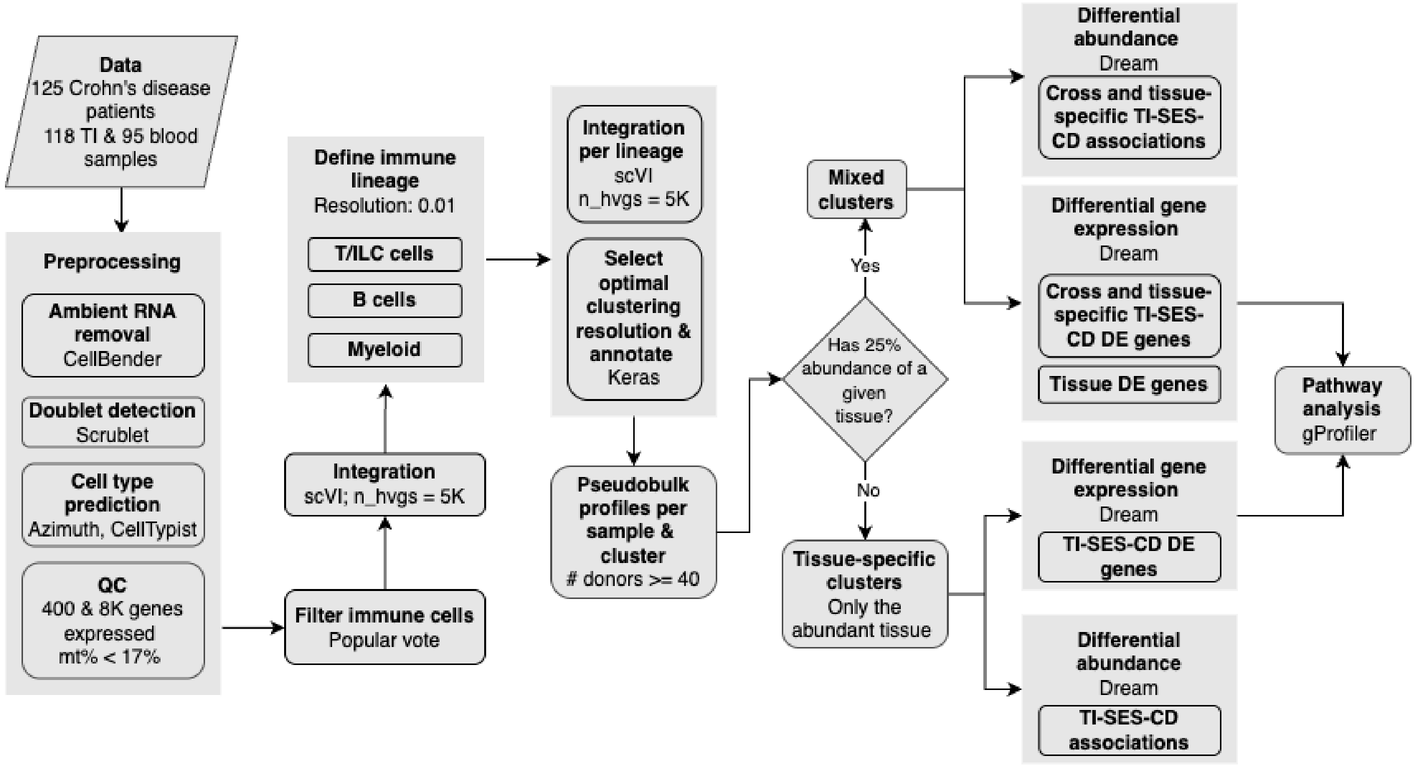


**Supplementary Fig. 1. Overview of the analysis.** Workflow of the data processing and analysis pipeline used to identify tissue-specific and cross-tissue associations with mucosal inflammation, as measured by the Simple Endoscopic Scores for Crohn Disease at the terminal ileum (TI-SES-CD). The workflow includes preprocessing, cell type annotation, lineage-specific integration and clustering, pseudobulk profiling and differential abundance and expression analyses. Mixed and tissue-specific clusters were analyzed separately, followed by pathway enrichment using gProfiler.

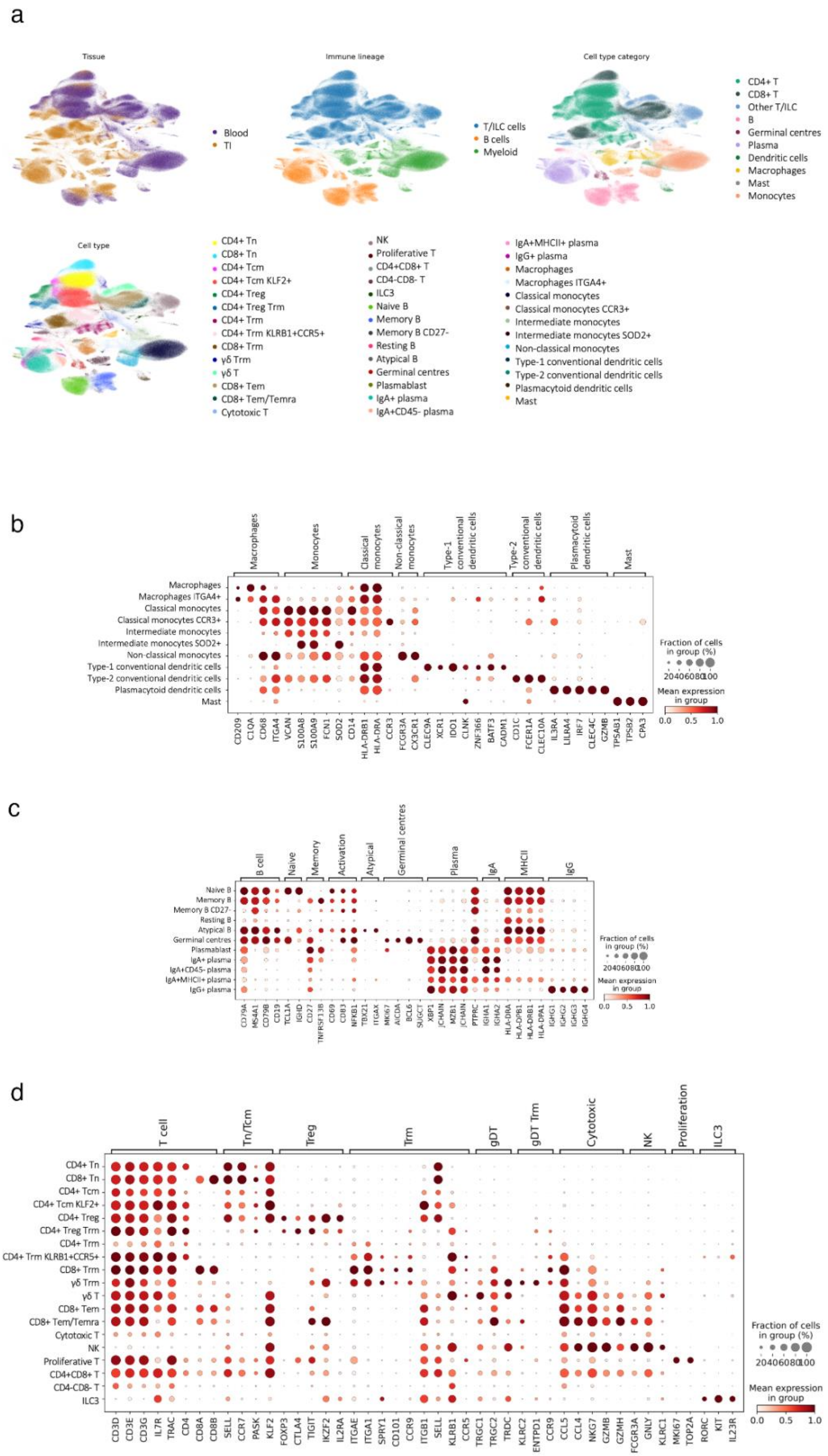

a
Tissue
Blood
TI
Immune lineage
T/ILC cells
B cells
Myeloid
Cell type category
CD4+ T
CD8+ T
Other T/ILC
B
Germinal centres
Plasma
Dendritic cells
Macrophages
Mast
Monocytes
Cell type
CD4+ Tn
CD8+ Tn
CD4+ Tcm
CD4+ Tcm KLF2+
CD4+ Treg
CD4+ Treg Trm
CD4+ Trm
CD4+ Trm KLRB1+CCR5+
CD8+ Trm
γδ Trm
γδ T
CD8+ Tem
CD8+ Tem/Temra
Cytotoxic T
NK
Proliferative T
CD4+CD8+ T
CD4-CD8- T
ILC3
Naive B
Memory B
Memory B CD27-
Resting B
Atypical B
Germinal centres
Plasmablast
IgA+ plasma
IgA+CD45- plasma
IgA+MHCII+ plasma
IgG+ plasma
Macrophages
Macrophages ITGA4+
Classical monocytes
Classical monocytes CCR3+
Intermediate monocytes
Intermediate monocytes SOD2+
Non-classical monocytes
Type-1 conventional dendritic cells
Type-2 conventional dendritic cells
Plasmacytoid dendritic cells
Mast
b
Macrophages
Monocytes
Classical monocytes
Non-classical monocytes
Type-1 conventional dendritic cells
Type-2 conventional dendritic cells
Plasmacytoid dendritic cells
Mast
Macrophages
Macrophages ITGA4+
Classical monocytes
Classical monocytes CCR3+
Intermediate monocytes
Intermediate monocytes SOD2+
Non-classical monocytes
Type-1 conventional dendritic cells
Type-2 conventional dendritic cells
Plasmacytoid dendritic cells
Mast
CD209 C1QA CD68 ITGA4 VCAN S100A8 S100A9 FCN1 SOD2 CD14 HLA-DRB1 HLA-DRA CCR3 FCGR3A CX3CR1 CLEC9A XCR1 IDO1 CLNK ZNF366 BATF3 CADM1 CD1C FCER1A CLEC10A IL3RA LILRA4 IRF7 CLEC4C GZMB TPSAB1 TPSB2 CPA3
Fraction of cells in group (%)
20 40 60 80 100
Mean expression in group
0.0 0.5 1.0
c
B cell
Naive
Memory
Activation
Atypical
Germinal centres
Plasma
IgA
MHCII
IgG
Naive B
Memory B
Memory B CD27-
Resting B
Atypical B
Germinal centres
Plasmablast
IgA+ plasma
IgA+CD45- plasma
IgA+MHCII+ plasma
IgG+ plasma
CD79A MS4A1 CD79B CD19 TCL1A IGHD CD27 TNFRSF13B CD69 CD83 NFKB1 TBX21 ITGAX MKI67 AICDA BCL6 SUGCT XBP1 JCHAIN MZB1 JCHAIN PTPRC IGHA1 IGHA2 HLA-DRA HLA-DPB1 HLA-DRB1 HLA-DPA1 IGHG1 IGHG2 IGHG3 IGHG4
Fraction of cells in group (%)
20 40 60 80 100
Mean expression in group
0.0 0.5 1.0
d
T cell
Tn/Tcm
Treg
Trm
gDT
gDT Trm
Cytotoxic
NK
Proliferation
ILC3
CD4+ Tn
CD8+ Tn
CD4+ Tcm
CD4+ Tcm KLF2+
CD4+ Treg
CD4+ Treg Trm
CD4+ Trm
CD4+ Trm KLRB1+CCR5+
CD8+ Trm
γδ Trm
γδ T
CD8+ Tem
CD8+ Tem/Temra
Cytotoxic T
NK
Proliferative T
CD4+CD8+ T
CD4-CD8- T
ILC3
CD3D CD3E CD3G IL7R TRAC CD4 CD8A CD8B SELL CCR7 PASK KLF2 FOXP3 CTLA4 TIGIT IKZF2 IL2RA ITGAE ITGA1 SPRY1 CD101 CCR9 ITGB1 SELL KLRB1 CCR5 TRGC1 TRGC2 TRDC KLRC2 ENTPD1 CCR9 CCL5 CCL4 NKG7 GZMB GZMH FCGR3A GNLY KLRC1 MKI67 TOP2A RORC KIT IL23R
Fraction of cells in group (%)
20 40 60 80 100
Mean expression in group
0.0 0.5 1.0

**Supplementary Fig. 2. Cell type annotation of mucosal and circulating immune cells in Crohn's disease.** a, Uniform manifold approximation and projection (UMAP) of immune cells from the terminal ileum (TI) and blood, colored by tissue of origin (top left) and cell type annotations at different clustering levels (immune lineage, cell type category and cell type; top right and bottom panels). b-d, Dot plots showing the scaled mean expression of canonical marker genes used for annotation of (b) myeloid cells, (c) B cells and (d) T cells. Dot size indicates the fraction of cells within each cluster expressing the gene; color intensity represents the scaled mean expression within each gene.

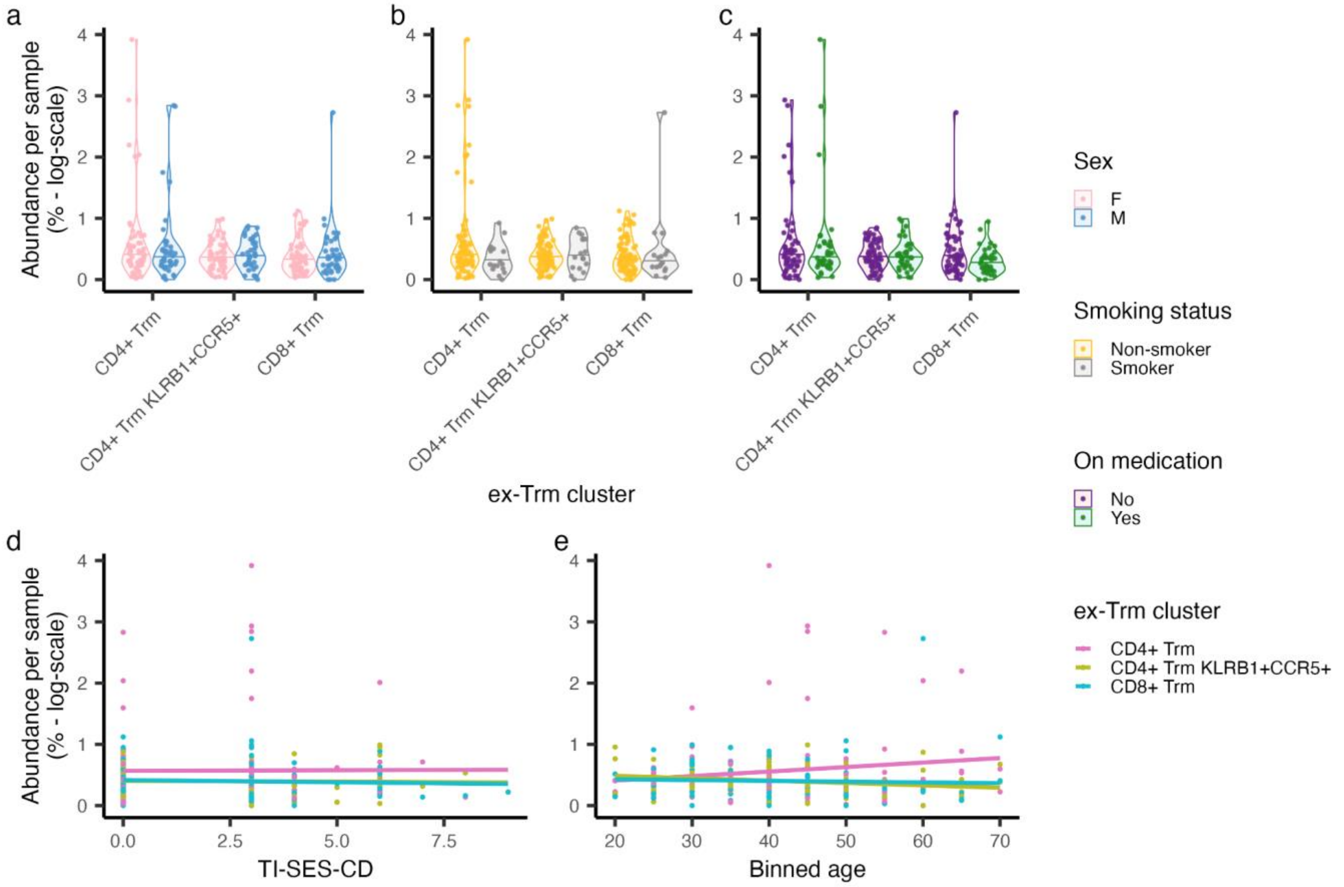


**Supplementary Fig 3. Associations between ex-Trm abundance and clinical or demographic variables.** a-c, Violin plots showing per-sample abundance (log-transformed) of ex-Trm clusters stratified by (a) sex, (b) smoking status and (c) medication status. d-e, Scatter plots showing the relationship between ex-Trm abundance and (d) the patient's Simple Endoscopic Scores for Crohn Disease at the terminal ileum (TI-SES-CD) or (e) age (binned). Statistical associations from beta regression models are reported in Supplementary Table 3.

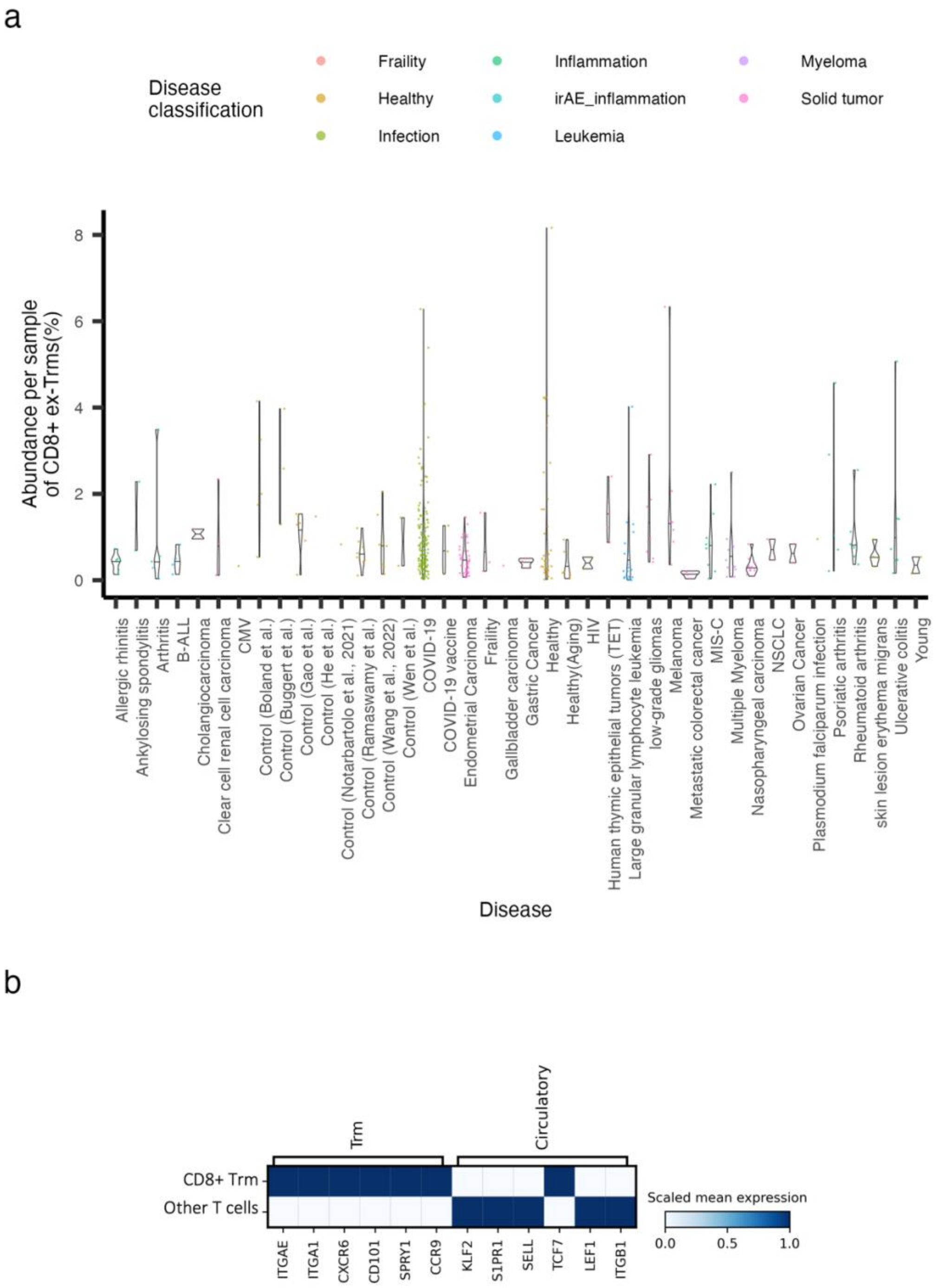


**Supplementary Fig. 4. Detection of ex-Trms across independent blood single-cell datasets.** a, Abundance of cells confidently predicted as $CD8^+$ ex-Trms (confidence > 0.5) in peripheral blood mononuclear cell (PBMC) datasets from a meta-analysis of $CD8^+$ T cells. Cell abundance is colored according to disease classification reported in the original publication. b, Heatmap showing the scaled mean expression of tissue-resident memory (Trm) and circulatory markers in predicted ex-Trms compared to the other circulating $CD8^+$ T cells.

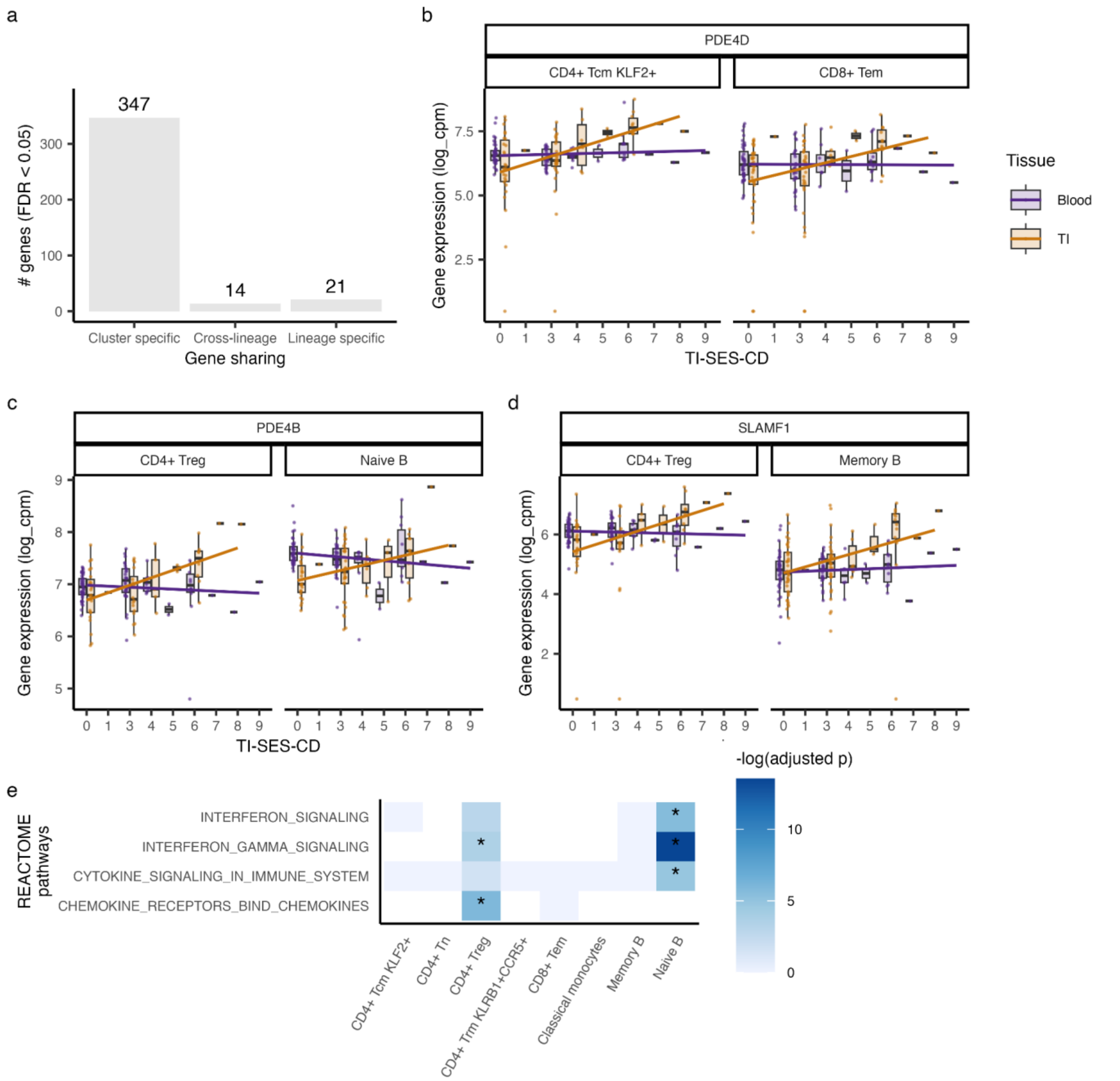


**Supplementary Fig. 5. Tissue-specific inflammation-associated gene sharing and pathway enrichment.** a, Bar plot showing the number of tissue-specific differentially expressed (DE) genes associated with the Simple Endoscopic Score for Crohn's Disease at the terminal ileum (TI-SES-CD). Genes are grouped by whether they are specific to individual clusters, shared within immune lineages (T/ILC cells, B cells or myeloid), or shared across lineages. b-d, Boxplots showing expression patterns of selected tissue-specific DE genes detected in multiple clusters. Lines represent linear regression fits across the TI-SES-CD axis, separated by tissue (TI or blood). e, Heatmap showing significant REACTOME pathways enriched among tissue-specific TI-SES-CD DE genes across clusters. Asterisks indicate significant associations (adjusted $p < 0.05$).

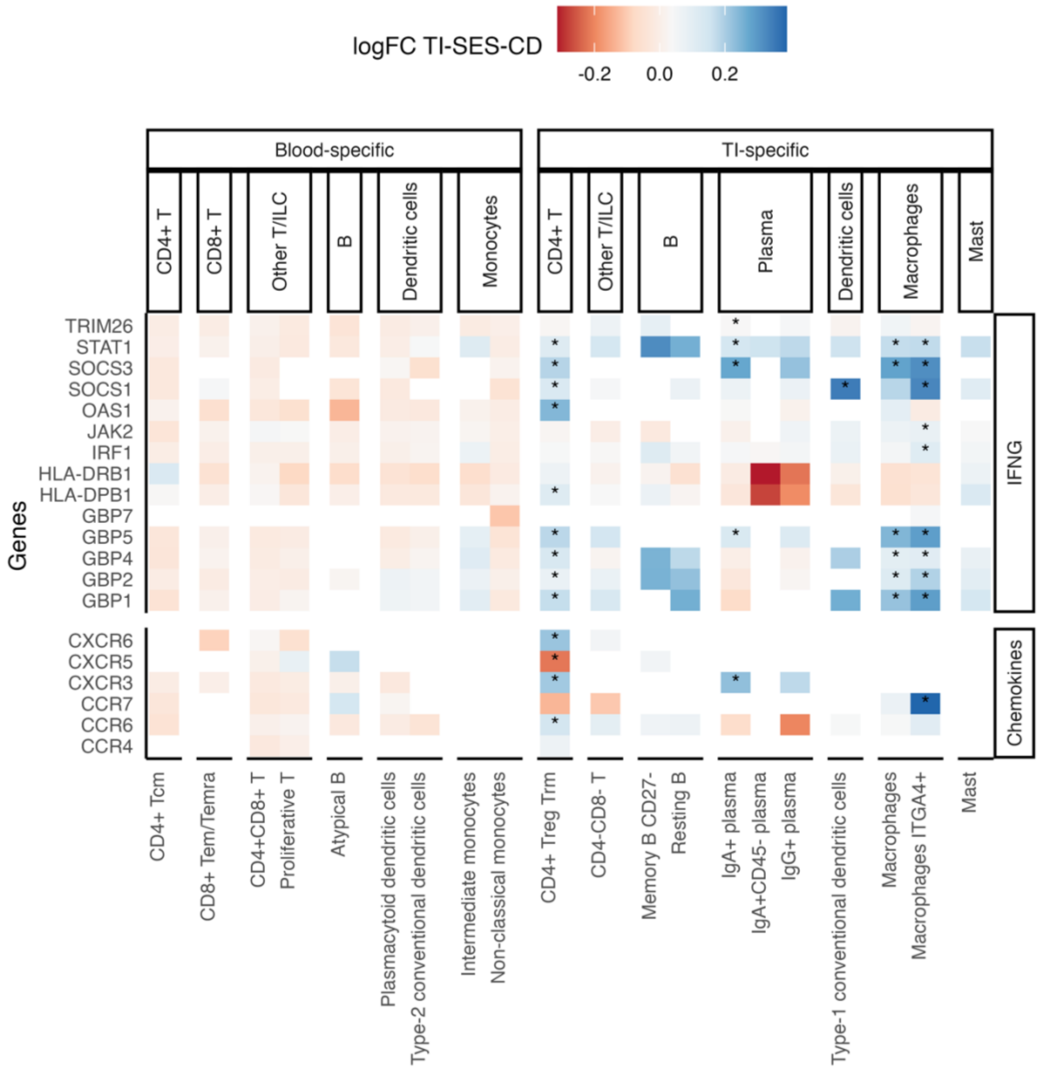


**Supplementary Fig. 6. Tissue-specific associations of IFNγ and chemokine signaling genes with mucosal inflammation.** Heatmap showing log fold-change (logFC) values between gene expression and the Simple Endoscopic Score for Crohn's Disease at the terminal ileum (TI-SES-CD) across blood-specific and TI-specific immune cell clusters. Genes were selected from REACTOME pathway enrichment analysis of tissue-specific TI-SES-CD differentially expressed genes and include those involved in interferon gamma (IFNγ) signaling and chemokine signaling. Asterisks indicate significant associations (FDR < 0.05).

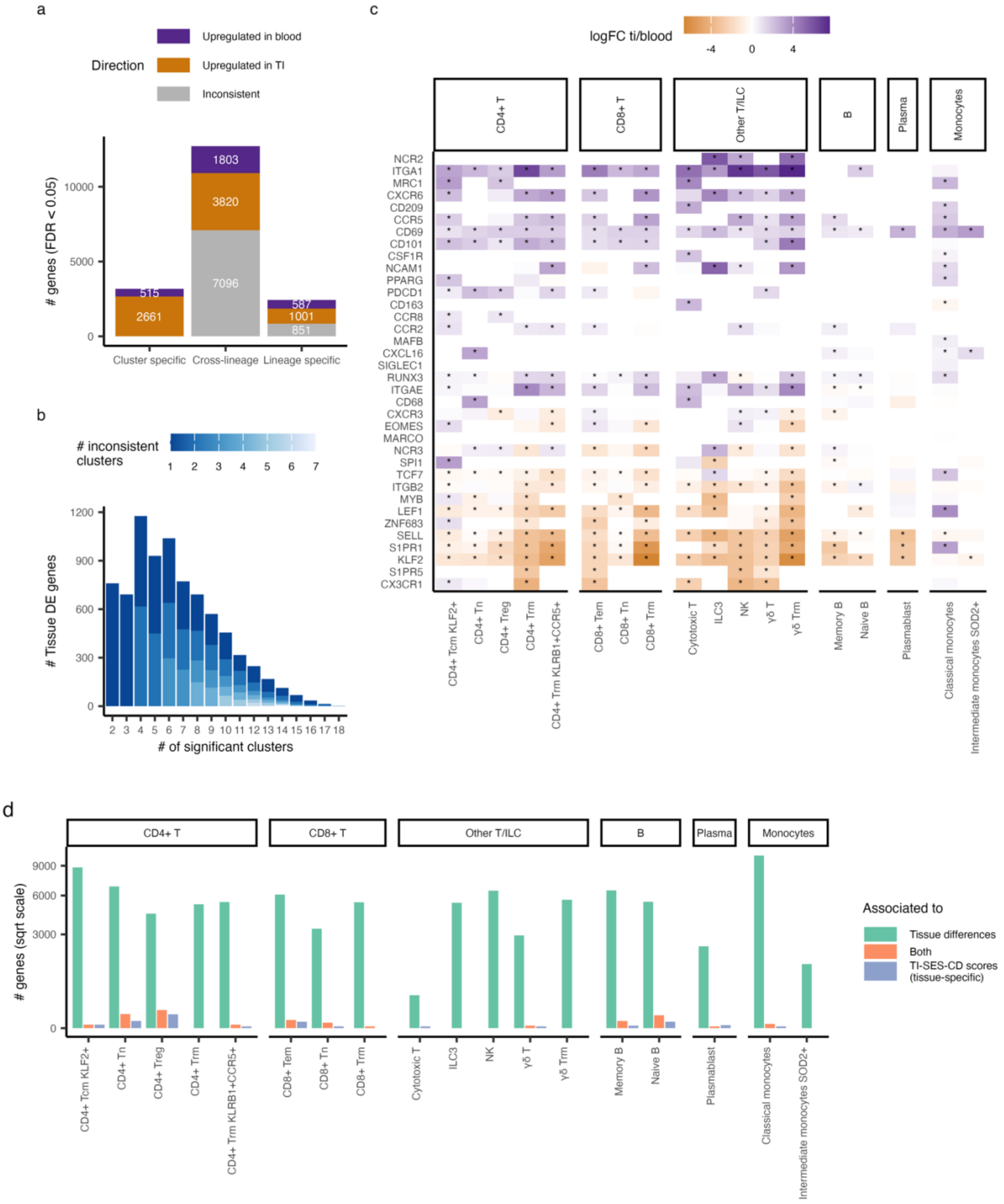

a
Direction
Upregulated in blood
Upregulated in TI
Inconsistent
### genes (FDR < 0.05)
1803
3820
7096
515
2661
587
1001
851
Cluster specific
Cross-lineage
Lineage specific
b
### inconsistent clusters
### Tissue DE genes
### of significant clusters
c
logFC ti/blood
CD4+ T
CD8+ T
Other T/ILC
B
Plasma
Monocytes
NCR2
ITGA1
MRC1
CXCR6
CD209
CCR5
CD69
CD101
CSF1R
NCAM1
PPARG
PDCD1
CD163
CCR8
CCR2
MAFB
CXCL16
SIGLEC1
RUNX3
ITGAE
CD68
CXCR3
EOMES
MARCO
NCR3
SPI1
TCF7
ITGB2
MYB
LEF1
ZNF683
SELL
S1PR1
KLF2
S1PR5
CX3CR1
CD4+ Tcm KLF2+
CD4+ Tn
CD4+ Treg
CD4+ Trm
CD4+ Trm KLRB1+CCR5+
CD8+ Tem
CD8+ Tn
CD8+ Trm
Cytotoxic T
ILC3
NK
γδ T
γδ Trm
Memory B
Naive B
Plasmablast
Classical monocytes
Intermediate monocytes SOD2+
d
### genes (sqrt scale)
Associated to
Tissue differences
Both
TI-SES-CD scores (tissue-specific)

**Supplementary Fig. 7. Sharing and specificity of tissue-associated differentially expressed genes.** a, Stacked bar plot showing the number of tissue-specific differentially expressed (DE) genes that are cluster-specific, shared within immune lineages (T/ILC cells, B cells or myeloid), or shared across lineages. Bar colors indicate directionality of expression: upregulated in blood, upregulated in the terminal ileum (TI) or inconsistent directionality across mixed clusters. b, Stacked bar plot showing the number of cross-lineage DE genes with inconsistent directionality of effect, stratified by the number of clusters in which the gene was significantly different across tissues. c, Heatmap showing log fold-change (logFC) values for curated gene markers associated with tissue-resident immune cells across mixed clusters. Asterisks denote significant expression differences between blood and TI (FDR $< 0.05$). d, Bar plot showing the number of genes with tissue-specific differences, associations with the Simple Endoscopic Score for Crohn's Disease at the terminal ileum (TI-SES-CD), or both, across mixed clusters.

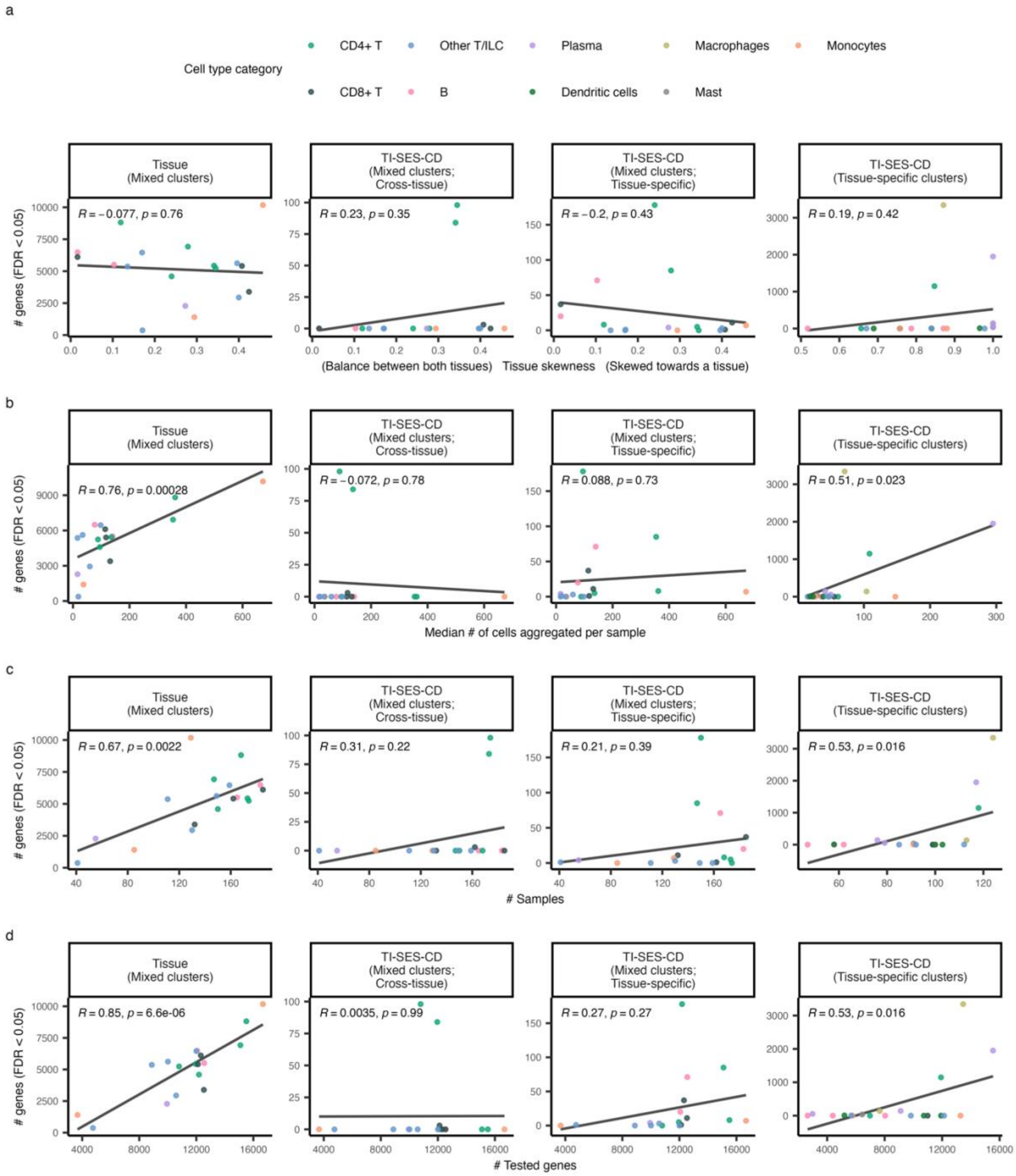


**Supplementary Fig. 8. Influence of cluster characteristics on differential gene expression power.** a-d Correlation analyses between the number of differentially expressed (DE) genes and key cluster characteristics, including tissue skewness that measures the deviation from a 1:1 blood-to-gut sample ratio (a), median number of cells aggregated per sample (b), total number of samples (c) and total number of genes tested (d). Each panel shows correlations stratified by the covariate used in the differential gene expression analysis: tissue-associated or inflammation-associated, as measured by the Simple Endoscopic Score for Crohn's Disease at the terminal ileum (TI-SES-CD). Inflammation-associated DE genes from mixed clusters are further divided into cross-tissue and tissue specific effects. Points are colored by the cell type category. Pearson correlation coefficients (R) and associated p-values are shown in each panel.

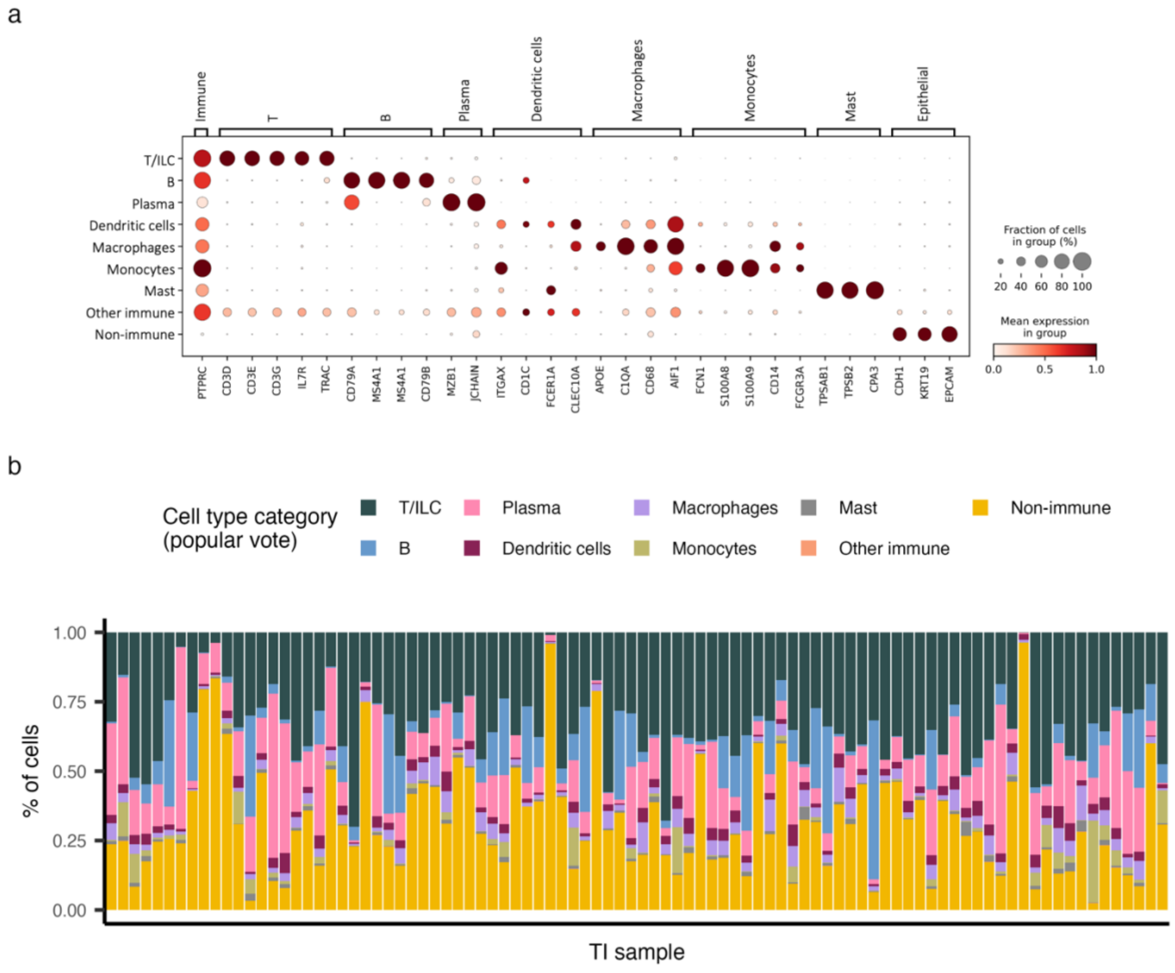


**Supplementary Fig. 9. Annotation of non-immune cells in terminal ileum samples.** a, Dot plot showing scaled mean expression of canonical marker genes across harmonized cell type categories in terminal ileum (TI) samples. Dot size indicates the fraction of cells expressing each gene and color intensity reflects the scaled mean expression within each gene. Cell type annotations were assigned to individual cells by majority vote across three independent gut single-cell atlases. b, Stacked bar plot showing the proportion of each harmonized cell type category across individual TI samples.

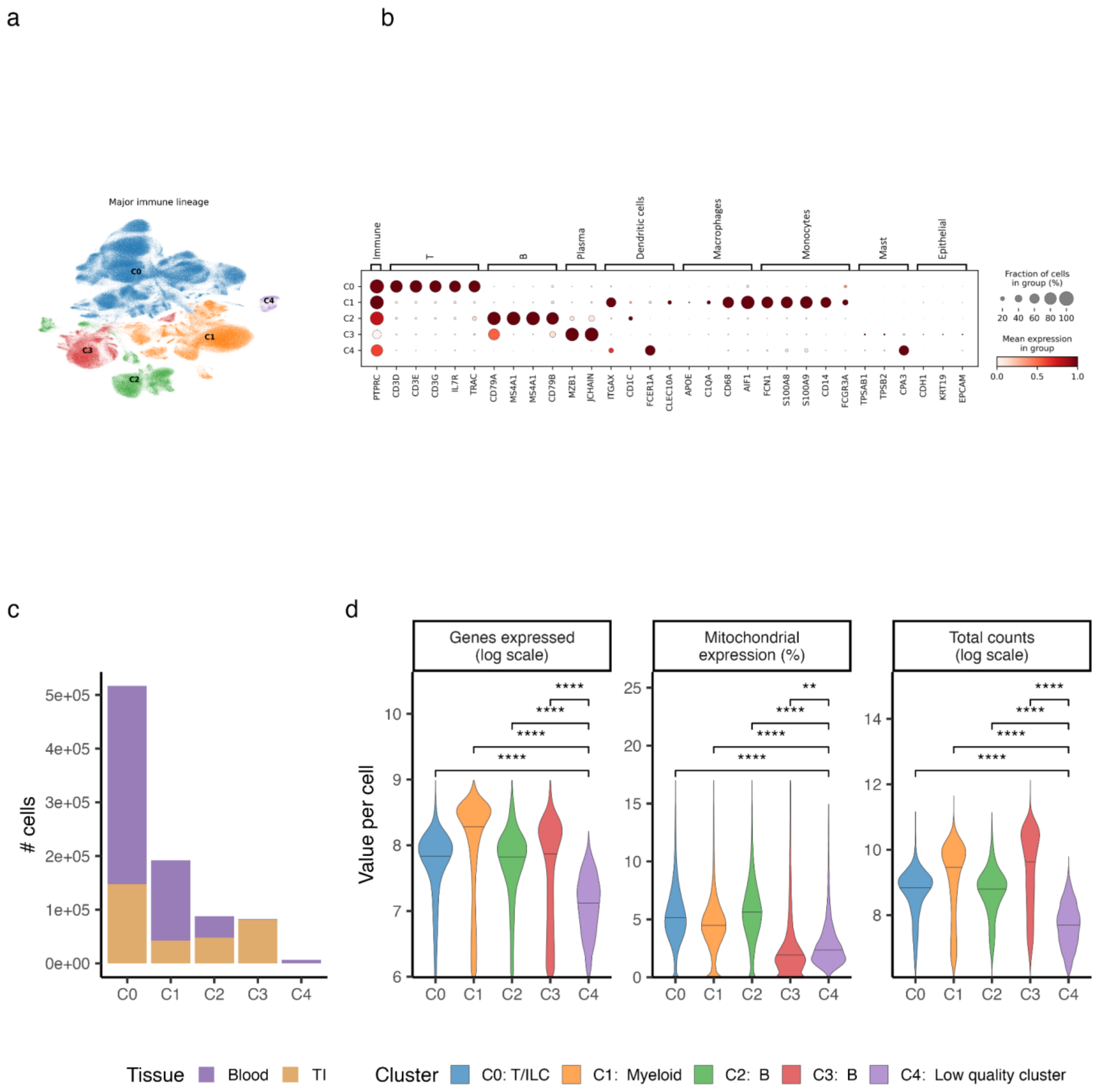


**Supplementary Fig. 10 Annotation of immune lineages across mucosal and blood compartments.** a, Uniform manifold approximation and projection (UMAP) of immune cells from terminal ileum (TI) and blood, colored by Leiden clustering. b, Dot plot showing the scaled mean expression of canonical marker genes across clusters. c, Bar plot showing the number cells per cluster, colored by tissue of origin (blood or TI). d, Violin plots displaying per-cell quality control metrics across clusters, including the number of genes expressed, mitochondrial gene expression percentage and total UMI counts. P-values are derived from t-tests comparing cluster C4 to each other cluster.

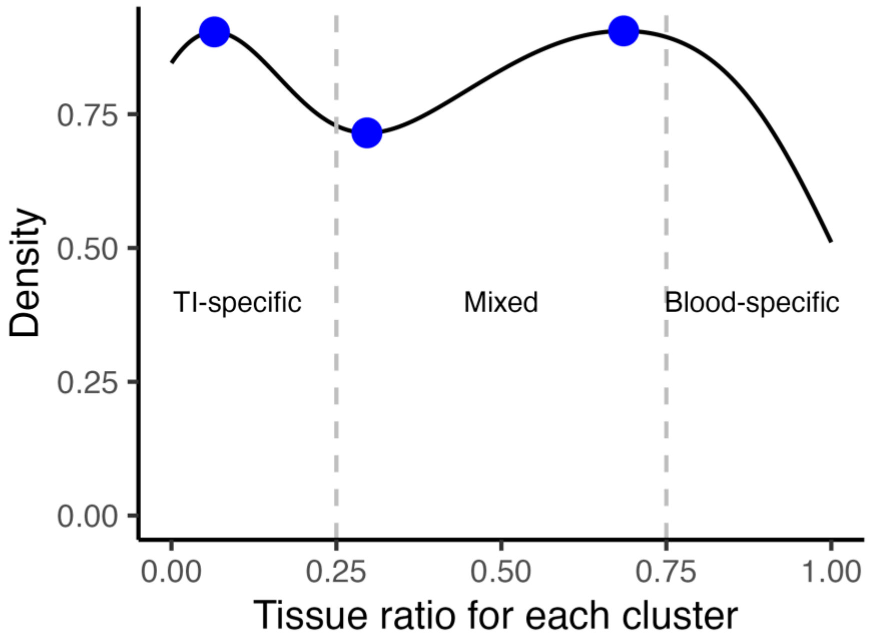


**Supplementary Fig. 11. Classification of clusters based on tissue composition.** Density plot showing the tissue ratios (blood versus terminal ileum, TI) across all 41 clusters. The x-axis represents the proportion of samples from blood within each cluster (0 = all TI, 1 = all blood). Vertical dashed lines indicate threshold values used to classify clusters as TI-specific, mixed or blood-specific. Blue dots represent local maxima and minima in the distribution, used to guide threshold selection.

### Supplementary Table legends

Supplementary Table 1. Clinical characteristics of Crohn's disease patients included in the atlas of mucosal and circulating immune cells.

Supplementary Table 2. Cell type annotation and label harmonization across immune and gut-related atlases. The first sheet summarizes lineage-level annotation and the remaining sheets detail lineage-specific annotation for T/ILC cells, B cells and myeloid cells. For each cluster and atlas, the two most represented external labels are reported, with the percentage of annotated cells indicated in parentheses.

Supplementary Table 3. Beta regression results assessing the relationship between per-sample abundance of each ex-Trm cluster and clinical covariates: sex, age, medication, mucosal inflammation, measured by the Simple Endoscopic Score for Crohn's Disease at the terminal ileum (TI-SES-CD) and smoking status.

Supplementary Table 4. Differential gene expression (DGE) analysis associating gene expression with mucosal inflammation (TI-SES-CD) across clusters. For mixed clusters, both tissue-specific (coefficient = ses_cd:is_ti) and cross-tissue (coefficient = ses_cd) associations are provided. For tissue-specific clusters, only the predominant tissue is tested (coefficient = ses_cd).

Supplementary Table 5. Follow-up DGE analysis on significant genes from Supplementary Table 4 to determine the direction of tissue-specific inflammatory effects. Separate models were run per cluster and tissue.

Supplementary Table 6. Differential abundance analysis correlating cluster proportions with the patient's TI-SES-CD score. For mixed clusters, both tissue-specific (coefficient = ses_cd:is_ti) and cross-tissue (coefficient = ses_cd) effects are shown; for tissue-specific clusters, only the predominant tissue was assessed.

Supplementary Table 7. REACTOME pathway enrichment for genes significantly associated with inflammation (from Supplementary Table 4). Results are stratified by positive, negative, and tissue-specific effects per cluster.

Supplementary Table 8. DGE results for tissue-associated transcriptional differences, independent of inflammation, across all mixed clusters.

Supplementary Table 9. REACTOME pathway enrichment for genes differentially expressed between blood and ileal samples (from Supplementary Table 8). Upregulated genes in each tissue are shown per cluster.

Supplementary Table 10. REACTOME pathway analysis of the intersection between inflammation-associated and tissue-associated genes for each cluster.
